# Interpretable machine learning and signal processing for automated reading and quality control of lateral flow tests for schistosomiasis

**DOI:** 10.1101/2025.10.01.25337079

**Authors:** Chris Ho, Christin Puthur, Betty Nabatte, Carson P. Moore, Theresia Abdoel, Rene Paulussen, Pafue Nganjimi, Pytsje T. Hoekstra, Narcis B. Kabatereine, Bumali Kawesa, John Odea, Ronald Bogere, Rosette Katushabe, Govert van Dam, Thomas F. Scherr, Goylette F. Chami

## Abstract

There is a lack of automated pipelines for diagnostic classification of point-of-care tests for neglected tropical diseases. Here we present an end-to-end automated pipeline for the analysis of point-of-care circulating cathodic antigen tests for schistosomiasis. We incorporated deep learning for cassette segmentation with signal processing. Automated classifications were compared to quantitative readings from calibrated antigen samples examined in lateral flow readers, and visual readings from highly trained field and senior technicians. The pipeline was evaluated for 3188 individuals within the SchistoTrack cohort in rural Uganda. Our quantitative classifications were on par with a lateral flow reader, and showed 86.6% sensitivity and 96.5% specificity with visual readings from a senior technician, which was an improvement on the visual readings from field technicians. Automated classifications were possible in as little as five minutes after test preparation for high antigen concentrations. We showed visual trace uncertainty can be resolved with signal processing, indicating visual traces should be classified as negative. Our pipeline will aid in advancing diagnostics to meet the World Health Organization target product profile for schistosomiasis, provide quantitative assessments for other diagnostics, enable large-scale surveillance in areas targeting elimination, and provide real-time quality control for diagnostics introduced into primary healthcare facilities.

## 1 Introduction

Lateral flow tests (LFTs) have changed the landscape of infectious disease diagnosis in resource-constrained settings by reducing the need to rely on presumptive clinical diagnosis and syndromic management.

Compared to formal laboratories requiring highly trained personnel and with lengthy result turnaround times, LFTs among other point-of-care (POC) tests do not require extensive equipment or expertise, are relatively inexpensive, and quick to administer [1]. LFTs are well established for malaria, human immunodeficiency virus (HIV), hepatitis B/C, severe acute respiratory syndrome coronavirus 2 (SARS-CoV-2), and tuberculosis as recommended for routine use by the World Health Organization (WHO) Essential Diagnostics List [2]. Yet, most LFTs fall short of meeting REASSURED diagnostic criteria that align with modern technological developments. REASSURED criteria incorporate real-time data analysis with ease of data collection in addition to the original ASSURED criteria of affordability, accessibility, and accuracy [3].

Machine learning (ML) has emerged recently as an invaluable tool for meeting REASSURED criteria. Integration of ML into LFT readings has provided methods to quickly denoise real-world data, reduce human error from subjective readings or untrained individuals, improve diagnostic classifications, and provide quantitative assessments of otherwise qualitative tests for improved sensitivity [4]. Yet, there remains a lack of interpretable end-to-end approaches that incorporate quality control and applications of ML for LFTs beyond HIV or SARS-CoV-2 [5] [6] [7].

A recent review [4] showed that existing ML approaches for LFTs focus on image processing and binary classification, using deep learning approaches. Human visual assessments are used as the training labels without validation through other diagnostics, curated samples with known antigen concentrations, or comparison to expected population-level infection prevalence. Blood and saliva are the most studied samples with less understanding of urine, despite known challenges in controlling differences in sample capillary flow [8]. Image libraries are used with minimal POC validation or most often no validation within the settings of clinical use [5]. Analyses thus far have focused on binary classification, despite ML providing the potential (if validated) for sensitive quantitative diagnoses [4].

Schistosomiasis LFTs, such as POC-CCA tests, are a prime example of a diagnostic without an automated end-to-end pipeline that could benefit from ML and other automated techniques to meet REASSURED criteria. 41 countries are estimated to be endemic for *Schistosoma mansoni* in sub-Saharan Africa (SSA) and require community-wide treatment in high (*>*50%) prevalence areas [9]. With recent cuts in international funding for mass drug administration (conducted without diagnosis [10]), and little-to-no availability of diagnostic tests or expertise within primary healthcare centres, there is an urgent need to improve the accessibility of schistosomiasis diagnostics [11]. Light microscopy remains the mainstay of diagnosis regardless of schistosome species [12]. The POC circulating cathodic antigen test (POC-CCA)–an immunochromatographic assay used with a urine sample [13]–for *S. mansoni* has shown promise in providing a more sensitive though less specific alternative to microscopy [14] [15]. Yet, POC-CCA still under performs compared to laboratory reference standards, though no true gold/reference standard exists [16] [17]. Common issues include the uncertainty around human visual classification of trace results [17] [14], batch-to-batch variation [18], shelf life variability, and unreliable results in low endemicity areas [19]. These issues remain due to the absence of real-time and remote quality control with a reliance on rarely accessed international reference laboratories that require cross-country shipment of patient samples [17].

We conducted a large-scale study of POC-CCA diagnoses within a real-world field setting for *S. mansoni* infections. 3188 individuals aged 5-90 were assessed within the SchistoTrack cohort with visual readings and an automated pipeline incorporating signal processing for result classification and deep learning for segmentation of POC-CCA cassettes. For comparison to signal processing, we also developed an end-to-end deep learning pipeline. Test-to-control ratios were validated against reference libraries with known schistosome antigen concentrations and visual readings from highly trained field and senior technicians. The aim of our study was to present a field-validated and interpretable end-to-end pipeline that could undertake image processing of POC-CCA tests, real-time quality control, and quantification of test results that is independent from visual labels.

## Results

### POC-CCA visual reading outcomes

There were 3188 participants with field-based valid POC-CCA tests and images available. Among the sampled 3188 participants, field technicians made the following visual assignments for POC-CCA results: 31.1% (992/3188) of participants were classified as negative, 16.6% (529/3188) as trace, and 52.3% (1667/3188) as positive. The prevalence was 68.9% (2196/3188) when considering trace as positive.

### Instance segmentation

Table 1 shows the training time, model size, and COCO mAP by class for each of the instance segmentation models. The reported mAP values include 95% CIs calculated by bootstrapping the test set. The best performing model in terms of cassette membrane mAP was the Mask R-CNN ResNet-50-FPN model trained on both the membrane and buffer well. This model achieved superior membrane mAP than both of the membrane-only models, including the larger ResNeXt model despite the ResNet being less computationally expensive to train.

**Table 1:**
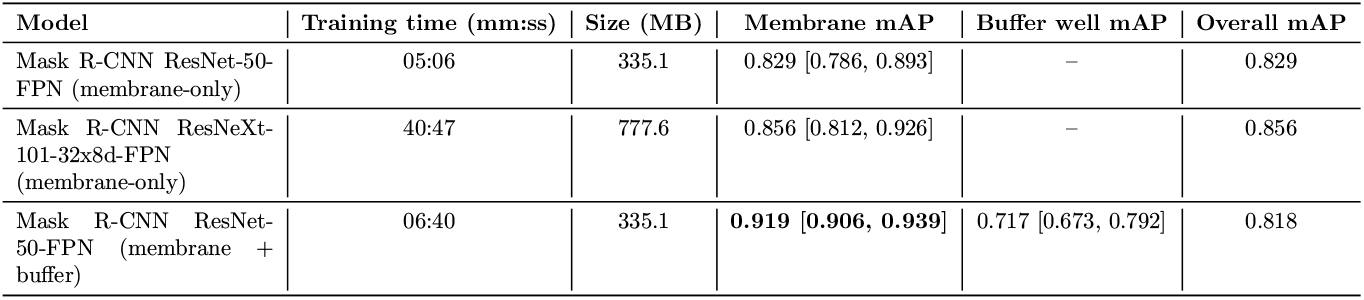
Segmentation results for deep learning models. The models were trained on 258 images from the training set of the field POC-CCA images for 12 epochs.

| Model | Training time (mm:ss) | Size (MB) | Membrane mAP | Buffer well mAP | Overall mAP |
| --- | --- | --- | --- | --- | --- |
| Mask R-CNN ResNet-50-FPN (membrane-only) | 05:06 | 335.1 | 0.829 [0.786, 0.893] | – | 0.829 |
| Mask R-CNN ResNeXt-101-32x8d-FPN (membrane-only) | 40:47 | 777.6 | 0.856 [0.812, 0.926] | – | 0.856 |
| Mask R-CNN ResNet-50-FPN (membrane + buffer) | 06:40 | 335.1 | **0.919 [0.906, 0.939]** | 0.717 [0.673, 0.792] | 0.818 |

### Accuracy for the AWA-TCA calibrated sample reference libraries

Figure 1 displays the results of the image analysis pipeline applied to the reference libraries as compared to the LFR results. The mean test-to-control line signal ratio obtained by image analysis of the reference library from the industry lab fell within the range of the LFR measurements for all calibrated samples except for CCA concentrations 120 and 240 ng/mL. The image analysis outcomes of the reference library from the government lab at the Uganda Ministry of Health slightly underestimated the test-to-control ratio compared to the LFR, but importantly displayed the same trend for test-to-control ratios as the LFR results. Treating the LFR as ground-truth, the mean absolute percentage error across all calibrated samples was 38.5% and 13.4% for the government lab and industry lab, respectively.

**Figure 1:**
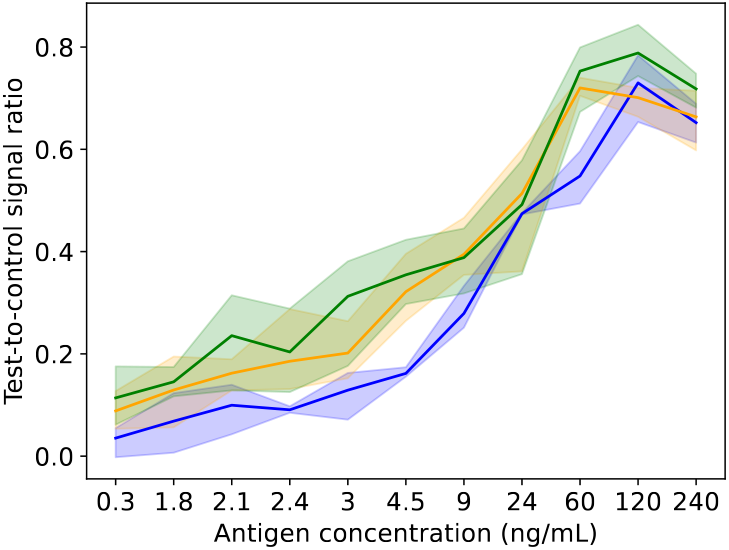
Pipeline performance on reference libraries. A comparison of image analysis outcomes is shown for the government lab at the Uganda Ministry of Health (blue) and the industry lab at MondialDx (yellow) with lateral flow readings from the research lab at Vanderbilt University (green). The graph plots the test-to-control ratio for the 11 calibrated samples examined. The shading shows the range of the triplicate readings, and the solid line is the mean of the three replicates.

### Image analysis outcomes for field images

The full runtime of the image analysis pipeline on 3188 images took 7 minutes 35 seconds (0.143s/image) on a V100 GPU. Factoring in one minute of field preparation of the cassette, the entire process to diagnosis took 8 minutes and 35 seconds. From the image analysis pipeline, using a threshold of 0.15 for the test-to-control ratio, 47% (1497/3188) of participants were classified as positive. The test-to-control ratio was nearly perfectly correlated with the test line signal (Spearman’s *ρ* = 0.96, *p <* 0.05), and only weakly correlated with the control line signal (Spearman’s *ρ* = 0.22, *p <* 0.05), indicating that the test signal dominates the ratio and no apparent bias by using a test-to-control ratio (Supplementary Figure S1). The median control signal intensity was slightly higher for positive linescan results (median 215.15, interquartile range (IQR) 167.12-296.02), and the difference was statistically significant (Wilcoxon rank-sum test *p <* 0.05; Supplementary Figure S1). However, this difference was small relative to the variation in test signal intensities, again indicating that variation in the control signal did not substantially bias the classification results.

Figure 2 shows the distribution of the test-to-control ratio for 3186 field POC-CCA images, overall and categorised by visual readings by field technicians. The test-to-control ratios for two of the 3188 field images with outlier values of 1.87 and 7.27 were excluded from the plots. The median test and control signals were 24.9 and 200.4, respectively (Supplementary Figure S2(a)), and the median test-to-control ratio was 0.13 overall. For binary visual classifications, the median test-to-control ratio was over nine times higher in images read as positive compared to those read as negative (0.27 vs 0.03 when trace was considered positive, Mann-Whitney U = 2043113, p *<* 0.05; 0.39 vs 0.04 when trace was considered negative, Mann-Whitney U = 2472332, p *<* 0.05; Figures 2(c), (d)). The median test-to-control ratios across the visual classifications of negative, trace, and positive were 0.03, 0.07, and 0.39, respectively (Kruskal-Wallis H=2250.3, p*<*0.05). The linescans indicated that visual traces were most likely negatives. All of the test-to-control ratios of visual trace results fell within two standard deviations of the test-to-control ratio distribution of visual negative results. Across all categories of visual results from the field, the lognormal distribution consistently had the lowest Bayesian Information Criterion (BIC) compared to the gamma and exponential distributions, indicating the best fit to the test-to-control ratio. BIC values for overall results and each visual assignment category are provided in Supplementary Table S2.

**Figure 2:**
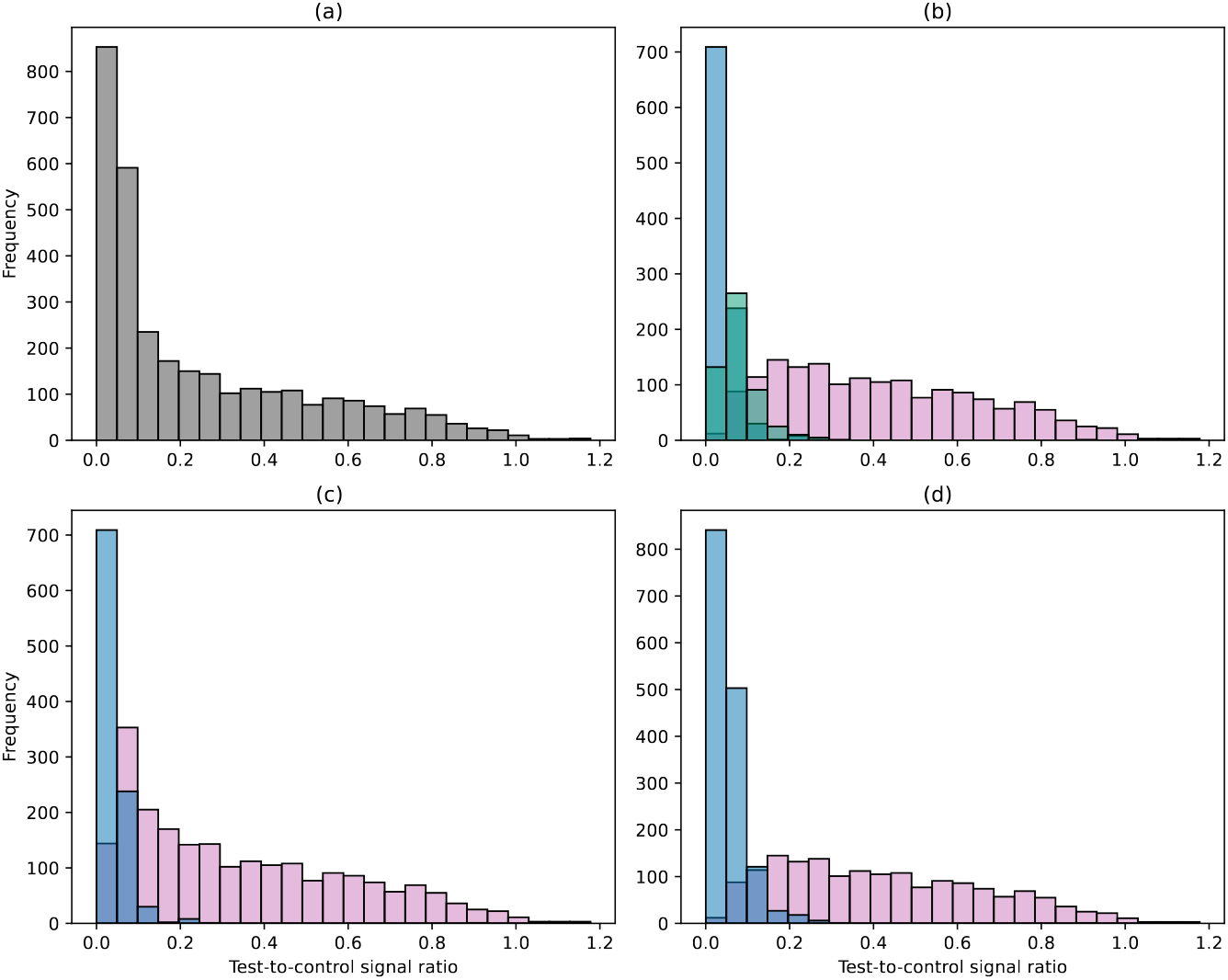
Distribution of test-to-control ratios. Results for 3186 field images are shown after removal of two results with outlier values of 1.87 and 7.27 from 3188. (a) Overall distribution, (b) Distribution by positive (pink), trace (green), and negative (blue), (c) Distribution by positive (pink) and negative (blue) where trace is considered positive, and (d) Distribution when trace is considered negative.

As expected in the design of the POC-CCA tests, the median test line signal across all image analysis outcomes was significantly lower than the median control line signal overall as well as for each visual assignment category (Supplementary Table S1; Supplementary Figure S2).

### Comparisons of test-to-control ratio and intensity distributions

The distributions of test line, control line, and test-to-control ratios showed clear separation across the negative, trace, and positive visual readings assigned by field technicians (Figure 2(b), Supplementary Figures S2(b)-(d)). For the test-to-control ratio, the distribution peak shifted progressively rightward from negative to trace to positive, mirroring the visual classification order. The test line signal distribution followed a similar trend, whereas the control line distribution exhibited only a slight rightward shift from negative to positive. All findings for the test-to-control ratio, test line, and control line distributions against the visual reading assignment remained robust when examined for the subset of field images used for senior technician rereading (Supplementary Figures S3(b), S4(b)-(d)). The distributions of test-to-control ratios, test lines, and control lines for the subset also remained similar against the visual assignments provided by the senior technician (Supplementary Figures S5(a), S6).

### Agreement with visual readings of field images

The confusion matrices shown in Figure 3 present the agreement between the visual readings by field technicians and the image analysis line-scans. When trace was considered positive, the agreement was 77.1% (2459/3188), with a sensitivity of 67.5% (1482/2196) and a specificity of 98.5% (977/992); whereas when trace was treated as negative, agreement rose to 91.3% (2912/3188), with a sensitivity of 86.6% (1444/1667) and a specificity of 96.5% (1468/1521). Here the calculations of sensitivity and specificity considered field technician readings as visual ground truth.

**Figure 3:**
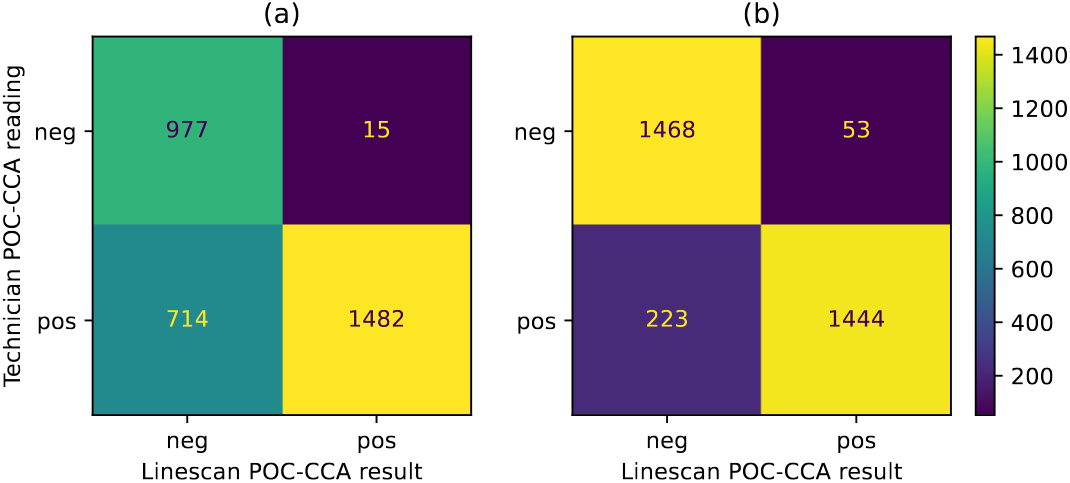
Agreement between visual readings for field technicians with image analysis outcomes. A threshold of 0.15 for the test-to-control ratio was considered positive for the image analysis outcome. (a) Trace readings were considered as positive results (b) Agreement with trace considered negative.

For the field images that were reread by a senior technician, the agreements between field technicians, senior technician rereadings, and image analysis outcomes are shown in the confusion matrices in Supplementary Figure S7. For the re-read images, the agreement between the senior technician and field technician was 76.4% (243/318) with trace readings as positive and 84% (267/318) with trace readings as negative. When agreement for the test-to-control ratios and visual readings was re-examined only for the subset of 318 images used for senior re-readings, the agreement between the field technician and image analysis outcomes was 74.5% (237/318) with trace readings as positive and 92.4% (294/318) with trace readings as negative. There was greater agreement between image analysis outcomes and senior technicians than between field technicians and senior technicians, irrespective of the coding of trace. Agreement between the senior technician and image analysis outcomes was 91.8% (292/318) with trace readings as positive and 88.4% (281/318) with trace readings as negative. The agreement percentages were similar across all three comparisons when 18 images with invalid time differences from the commercially-suggested reading time were excluded (Supplementary Figure S8). The distribution of test-to-control ratios and intensities for negative, trace, and positive was similar for the subset of 318 images when compared to the wider set of all 3188 field images (Supplementary Figures S3 S4). Considering the senior technician re-readings as the visual ground truth (Supplementary Figure S7), the linescan specificity improved from 51.6% to 92.9% compared with field technician readings when trace was considered positive, although sensitivity slightly decreased from 100% to 90.8%. When trace was considered negative, specificity increased from 73.7% to 81.4%, with sensitivity remaining high and relatively unchanged compared to field readings (100% vs 99.2%). Considering disagreements between field technician and image analysis outcomes by district, when trace was treated as positive, the proportion of disagreements was 20% (281/1404) in Pakwach, 22.3% (227/1020) in Buliisa, and 28.9% (221/764) in Mayuge, respectively, with a significant difference observed across the districts (*χ*^2^ = 22.6, *p <* 0.001). When trace was treated as negative, the proportion of disagreements was 8.4% (118/1404), 8.1% (83/1020), and 9.8% (75/764). These proportions did not differ significantly between districts (*χ*^2^ = 1.8, *p* = 0.415).

### Influence of deviations from the commercially-suggested reading time

Supplementary Figure S9 shows the time differences observed from the commercially-suggested reading time for 318 field POC-CCA images that were re-read by a senior technician. The median time difference was 2.7 minutes with an IQR of 1-7 minutes. Of these images, 65.7% (209/318) were taken within five minutes of the commercially-suggested reading time, with 9.1% (19/209) of these taken earlier, and 90.9% (190/209) taken later than the optimal time.

There was no overall association with the time the field image was taken and the test-to-control ratio extracted from the image analysis pipeline (Supplementary Figure S9(b). When the association was examined within the visual readings assigned by field technicians, the lack of association remained for subgroups of trace and positive as well as within WHO intensity categories of light, moderate, and heavy infection intensities from KK microscopy (Supplementary Figure S9(b-c)). Among only visual readings that already were classified as negative by field technicians, the larger the time difference, the lower the test-to-control ratio resulting in no change in the diagnostic classification. Results classified as no infection in the WHO KK categories also had a negative correlation with the time difference (Supplementary Figure S9(c). When considering the test signal values, significant negative correlation was observed only for visually negative results, and the WHO category of no infection (Supplementary Figure S10(a-b)). For the control signal, significant positive correlations with time difference were observed overall, for visual trace results, and for the moderate WHO intensity category (Supplementary Figure S10(c-d)). Considering agreement of visual results from the field with linescans, the median time difference for images with agreed results was not significantly different from the median time for results in disagreement (Supplementary Figure S11). The distribution of the test-to-control ratios, and the test and control signals for cases of disagreement in field and linescan results are shown in Supplementary Figure S12.

### Time-to-positivity

The test-to-control ratio over one-minute intervals from cassette preparation for four concentrations of calibrated samples are shown in Supplementary Figure S13. Second-order polynomials were fit to the scatter of each replicate. After 20 minutes, the test-to-control ratio was greater than 0.15–the threshold for a positive image analysis outcome–for all replicates of each sample. The time elapsed for crossing the test-to-control ratio threshold of 0.15 averaged over the three replicates was 23.7 minutes for the calibrated sample of CCA concentration 2.1 ng/mL, 16 minutes for the 3 ng/mL CCA sample, 14.3 minutes for 4.5ng/mL CCA, and 5 minutes for 9 ng/mL CCA. The unusual trend in one replicate (green) in Supplementary Figure S13 was driven by observing no change in the test line over time (Supplementary Figure S14). The control line signal reached its maximum across all four samples around 20 minutes; however, the peak signal in the test line was observed at 50 minutes for the calibrated sample with 2.1 ng/mL CCA, 33 minutes for 3 ng/mL CCA, 47.7 minutes for 4.5 ng/mL CCA, and 48 minutes for 9 ng/mL CCA, averaged over the three replicates (Supplementary Figure S14).

### Comparison to deep learning classification

Table 2 shows the sensitivity, specificity, and accuracy metrics for the deep learning classification experiments which used the same label source for training and testing, along with linescan-based results for comparison. The average total training times for ResNet50, MobileNetV2, and ViT were approximately 24, 26, and 47 minutes, respectively, when training on the larger field readings dataset, and around 7, 4, and 11 minutes on the smaller senior re-readings dataset. Results from a third experiment, where models trained on senior re-readings were applied to field-labeled images, are shown in Supplementary Table S3.

**Table 2:** Classifier performance comparisons.

| Classifier | Trace treatment | Trained and evaluated on field readings |  |  | Trained and evaluated on senior re-readings |  |  |
| --- | --- | --- | --- | --- | --- | --- | --- |
|  |  | Sensitivity | Specificity | Accuracy | Sensitivity | Specificity | Accuracy |
| ResNet50 | Positive | 0.930 | 0.859 | 0.908 | 0.680 | 0.957 | 0.812 |
|  |  | [0.905, 0.958] | [0.797, 0.913] | [0.883, 0.933] | [0.500, 0.857] | [0.857, 1.000] | [0.708, 0.917] |
| MobileNetV2 | Positive | 0.858 | 0.913 | 0.875 | 0.720 | 0.957 | 0.833 |
|  |  | [0.821, 0.891] | [0.865, 0.957] | [0.846, 0.902] | [0.548, 0.897] | [0.850, 1.000] | [0.729, 0.938] |
| ViT | Positive | 0.906 | 0.913 | 0.908 | 0.960 | 0.913 | 0.938 |
|  |  | [0.873, 0.938] | [0.864, 0.952] | [0.879, 0.933] | [0.864, 1.000] | [0.783, 1.000] | [0.854, 1.000] |
| Linescan | Positive | 0.694 | 0.980 | 0.783 | 0.920 | 0.957 | 0.938 |
|  |  | [0.638, 0.745] | [0.953, 1.000] | [0.741, 0.818] | [0.792, 1.000] | [0.864, 1.000] | [0.874, 1.000] |
| ResNet50 | Negative | 0.948 | 0.952 | 0.950 | 0.737 | 1.000 | 0.896 |
|  |  | [0.918, 0.973] | [0.923, 0.977] | [0.931, 0.969] | [0.533, 0.933] | [1.000, 1.000] | [0.812, 0.979] |
| MobileNetV2 | Negative | 0.952 | 0.948 | 0.950 | 0.842 | 1.000 | 0.938 |
|  |  | [0.925, 0.976] | [0.918, 0.975] | [0.929, 0.969] | [0.666, 1.000] | [1.000, 1.000] | [0.854, 1.000] |
| ViT | Negative | 0.940 | 0.952 | 0.946 | 0.947 | 0.897 | 0.917 |
|  |  | [0.910, 0.967] | [0.922, 0.978] | [0.925, 0.965] | [0.810, 1.000] | [0.769, 1.000] | [0.833, 0.979] |
| Linescan | Negative | 0.900 | 0.948 | 0.923 | 1.000 | 0.862 | 0.917 |
|  |  | [0.865, 0.936] | [0.918, 0.973] | [0.898, 0.946] | [1.000, 1.000] | [0.714, 0.968] | [0.833, 0.979] |

### Quantitative classifications

Figure 4 shows scatter plots of the test-to-control ratios and the signal intensities of the test lines against *log*(eggs per gram + 1). The test-to-control ratio (Spearman’s *ρ* = 0.59, *p <* 0.05) and test line signal intensity (Spearman’s *ρ* = 0.58, *p <* 0.05) were strongly positively correlated with eggs per gram. Strong correlations of test-to-control ratios and test signals were observed for field-assigned visual G-scores. Figure 5 shows a scatter plot of the test-to-control ratios (Spearman’s *ρ* = 0.91, *p <* 0.05) against G-score ratios, and a violin plot of signal intensities of the test lines (Spearman’s *ρ* = 0.93, *p <* 0.05) against the G-scores for test lines. When using G-scores to create a reference library, a similar sigmoidal shape was observed as with the other reference libraries from linescans and the LFR (Supplementary Figure S15).

**Figure 4:**
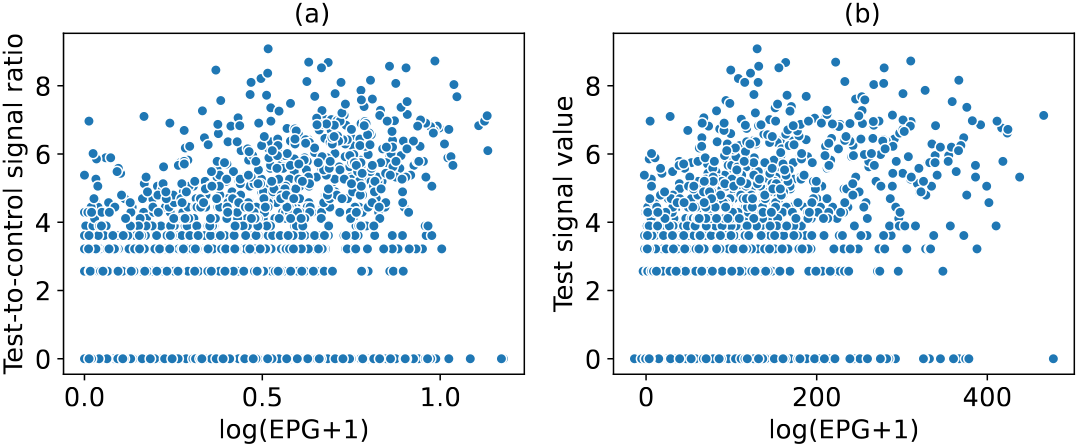
Association of linescan results with quantitative actual and proxy visual quantitative measurements. Association of eggs per gram with linescan results. Scatter plots of test-to-control ratios (a), and test signal values (b), against *log*(EPG + 1).

**Figure 5:**
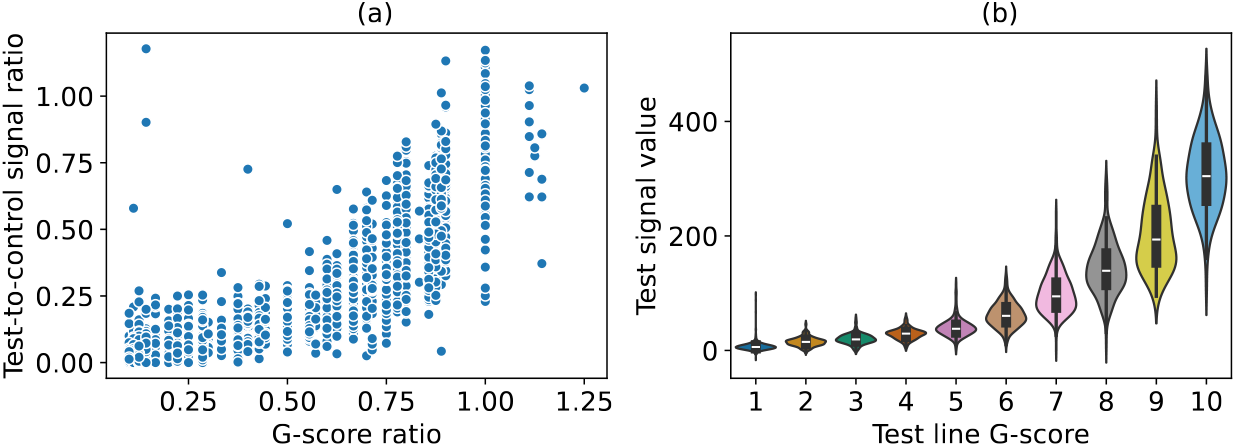
Association of G-scores with linescan results for field readings. Scatter plot of test-to-control ratios against G-score ratios (a), and violin plots of test signal values (b), against respective G-scores.

## Discussion

Improved diagnostics are needed for neglected tropical diseases, including schistosomiasis, to better target mass drug administration campaigns, improve field surveillance in low-transmission areas, and ultimately enable routine testing within primary healthcare facilities. Here we present a fully automated pipeline for accurate diagnostic classification and real-time quality control of schistosomiasis LFTs, focusing on the widely available POC-CCA test. We studied 3188 individuals in three rural districts of Uganda. Our automated pipeline performed on par with a LFR and showed high agreement with visual readings by a senior (expert) technician, showing also applicability for quantitative diagnostic classifications.

We developed an end-to-end automated pipeline for POC-CCA tests that performed well in a real-world field setting across three climatically diverse rural districts of Uganda. When automated diagnostic classifications were compared to four trained technicians who recorded visual readings at the POC for the participant, we observed agreement of up to 91.3% with 86.6% sensitivity and 96.5% specificity with trace considered negative. The field applicability was further supported in that agreement remained high despite a variety of temperatures from 28-35^°^C across study districts that may affect the analyte flow in the POC-CCA test. Disagreements between our automated pipeline and field technician readings were evenly distributed across the districts when trace was considered negative. The difference in minutes from when the image was taken and the optimal 20 minute reading timepoint also had no influence on the diagnostic classification. Importantly, we showed that images may be feasibly taken within five minutes of the commercially-suggested reading time in busy field settings like in SchistoTrack where 40 participants/patients are seen a day by a single technician. At the extreme, we would recommend no more than 20 minutes after the commercially-suggested reading window and ideally within 10 minutes for images to be used for automated classification, which is similar to the restrictions suggested by the POC-CCA test manufacturer for visual readings. The commercially-suggested reading window of 20 minutes was set for visual readings. In laboratory experiments, we found that a positive diagnostic classification can be determined in as little as 14 minutes for lower concentrations (e.g. 4.5 ng/mL antigen concentration), but in as little as five minutes for high concentrations (e.g. 9 ng/mL antigen concentration). Hence, our pipeline would be able to save time from the commercially suggested 20-minute reading window and is adaptable to the infection intensity of the individual (assuming antigen concentration correlates to fluke burden). A 6-15 minute savings in time is potentially the difference in seeing one versus two patients within a time-constrained and understaffed primary healthcare centre and may be a determining factor in the use of LFTs in emergency settings [20]. In practice, additional research is needed to determine whether these time savings can be achieved without apriori knowledge of participant infection intensity or constant observation by humans or camera/video of POC-CCA tests.

We introduced methods for remote quality control of POC-CCA tests. When new commercial batches of POC-CCA tests are produced, the quality of the batch relies on visual assessment of performance [21]. We showed that our pipeline is on par with a LFR, so can be used alongside calibrated samples with known antigen or other analyte concentrations to determine batch performance and expected saturation curves in order to measure batch-to-batch variability [22]. In SchistoTrack, POC-CCA tests were repeated up to three times to avoid the entry of invalid results given the need for POC diagnosis and treatment. However, in other settings where results may be sent and read remotely, our pipeline enables real-time quality control of individual POC-CCA tests by detecting and quantifying the control line signal. Depending on the test of interest, thresholds for a minimal control line signal could be set–whether to determine any presence or actual strength–and any values below that threshold could be deemed invalid. This procedure would be helpful for easily narrowing down when a repeated test is warranted. Most importantly, we demonstrated that our pipeline could also be used as a second senior reader if needed to check the quality of field classifications. Our automatic classifications agreed more with the senior (expert) technician than with field technicians. Using the senior technician as the reference standard, our pipeline improved specificity over the field technicians by 41.3% when trace was considered positive and by 7.7% when trace was considered negative, with only minimal reductions in sensitivity (9.2% and 0.8%, respectively). When scaled up to cover routine usage in primary healthcare facilities or large-scale field surveillance efforts in the context of elimination, these small changes in sensitivity and improvement may result in large cost savings [5].

Human uncertainty in POC-CCA test readings is common with barely visible treatment lines [4], but this uncertainty was resolved through automated classification. By looking at the signal intensity distribution of visually-classified trace ratios, we showed that the distribution did not significantly differ from the signal intensity distribution of negative tests. As KK microscopy is 100% specific, we also observed that 87.2% of trace results are negative by KK. From our study using highly trained field technicians, visual trace results observed in POC-CCA tests are recommended to be classified as negative results in future studies to improve test specificity. These results contrast other smaller-scale studies, mostly focused on school-aged children, where up to 50% of traces have been suggested as positive [23] [24]. Automated pipelines for image analyses of POC-CCA, due to the distribution of signal intensities, should consider trace as negative.

Although POC-CCA tests currently are only intended for qualitative readings, our pipeline enables future quantitative diagnostic classifications. We showed near perfect overlap with our pipeline and quantitative outputs using known antigen concentrations and a LFR. Conventionally, quantitative associations of POC-CCA have been correlated with KK microscopy where eggs from female flukes are counted in stool [21] [25]. We found positive correlations with the test-to-control ratios from our line scans and KK. Although this result provides support of quantification using POC-CCA, the results should be interpreted with caution due to the fundamental differences between the two diagnostics. POC-CCA detects fluke antigens whereas KK focuses on egg quantification. High concordance between POC-CCA and KK is not necessarily expected due to the presence of unpaired, immature, or reduced fecundity flukes. POC-CCA also has been shown to have poor specificity [19], resulting in positives that should not be quantified. Regardless, the pipeline developed here is indispensable for areas lacking calibrated LFRs and to maximise the value of forthcoming quantitative schistosomiasis LFTs such as the circulating anodic antigen (CAA) rapid diagnostic test [26]. Future field-based quantitative diagnostic classifications for schistosomiasis could improve test sensitivity to the limit required for monitoring the reduction in schistosome transmission and ultimately achieving focal elimination goals.

Our results show that linescan-based classification was comparable, and in some cases even exceeded performance of deep learning classification of the same set of segmented ROIs. While deep learning models are valued for their noise tolerance, they are dependent on human labels which restrict the validity of their results in the absence of a true gold standard. In contrast, the linescan approach is interpretable and produces quantitative results without the need for large labeled datasets. We also showed here that linescans can resolve trace ambiguity based on a consistent threshold for classification. Although the choice of a threshold might be considered a disadvantage over deep learning, the linescan allows reclassification if thresholds are updated and new criteria introduced, unlike in end-to-end deep learning classification.

Our pipeline has a number of policy implications relevant for meeting goals set in the WHO 2030 Roadmap for neglected tropical diseases (NTDs) [27]. Improved diagnostic performance is important for the design of future NTD control efforts where previous approaches to management and control (that do not rely on any diagnoses) such as mass drug administration are no longer sustainable options with limited medicine donations, participant fatigue, and reductions in international aid [28]. The improved sensitivity and specificity provided by our pipeline may aid in future surveillance efforts to progress towards regional elimination of schistosomiasis as well as substantial cost savings for national programmes. With integrated remote quality control, home test-and-treat [29] options are a possibility for schistosomiasis surveillance. Health system integration for schistosomiasis cannot rely on traditional microscopy, which for intestinal schistosomiasis, can take up to two days, two expert readers, and substantial resources to return a result. Automated analyses and quality control of POC-CCA tests could provide checks on shelf life of the tests, quick diagnoses for triage, and utilise any existing health centre staff regardless of training.

We proposed an end-to-end machine learning and signal processing pipeline with integrated quality control for POC-CCA tests. Automated approaches are becoming integral to ensure diagnostic performance in challenging, resource-constrained settings [30]. We anticipate that our pipeline will help to meet the WHO ideal target product profile [31] for schistosomiasis diagnostics that are quantitative, highly specific, low cost, high throughput, require no additional resources or maintenance, benefit from real-time connectivity for automated quality control, and can be used with little expertise.

## Methods

### Context and participant sampling

This study was nested in the ongoing community-based prospective SchistoTrack Cohort which assesses disease development related to intestinal schistosomiasis in rural Uganda [32]. SchistoTrack covers 52 villages across three districts of Pakwach, Buliisa, and Mayuge. The prevalence of *S. mansoni* ranged from 5-20% across individuals aged 5-90 years [33]. Using local council (village government) and neglected tropical disease (NTD) registers, 1952 households were randomly sampled across the 52 villages, with a maximum of 40 households per village. Within each household, one child aged 5-17 years and one adult aged 18+ years were selected by the household head and/or spouse to attend clinical assessments. This study was conducted during an annual follow-up of SchistoTrack in January-February 2023 where all participants attending that follow-up were analysed. Fieldwork was completed in the dry season and in open air locations at rural primary schools, churches, or similar community buildings where there was no electricity. In January 2023, the average temperature varied in the study districts from 35^°^C in Pakwach, 34^°^C in Buliisa, and 28^°^C in Mayuge [34].

### Sample and test preparation

Participants were provided a plastic sample pot of 100 ml with a secure twist on lid by village health team members and team surveyors with instructions to provide a fresh urine sample on the morning for which they will attend clinical assessments. A team of four laboratory technicians with 0-2 years of experience specifically reading POC-CCA tests, but everyone with at least one year of experience doing laboratory work within stringent SchistoTrack protocols and who had undergone a day of training with the POC-CCA tests were tasked with preparing urine samples and reading POC-CCA tests. For sample quality control, technicians conducted an initial inspection of the sample. They confirmed if the participant had provided enough sample (minimum of 20 ml) and if the sample was contaminated with any debris, dirt, or blood from menses. If the sample was not enough or contaminated, the participant was given a new sample pot and asked to provide a fresh sample on the same day.

ICT Diagnostics Schisto POC-CCA^®^ tests were used (batch number 221117133) [35]. Preparation of the POC-CCA tests began by labelling the cassette housing with a preassigned participant ID using a fine marker pen. The commercially-suggested reading time, as recommended by the manufacturer, of 20 minutes after test preparation was written onto the cassette. POC-CCA tests were then immediately prepared by using the commercially supplied straw droppers to place two drops of the urine into the sample well. Direct urine samples were used; no buffer solution was provided with the test. No more than five tests were performed at the same time.

### Field technician readings and image capture

At the commercially-suggested reading time, field technicians visually read and recorded the results. Tests were considered invalid if the control line was not visible to the technicians. Urine samples with an invalid test would be tested with new cassettes up to three times using the same urine sample before recording of an invalid result. There were no tests that were invalid in our study due to the repeated POC-CCA tests. The test line was classified by technicians by direct visual assessment (standard convention). Visual readings included three classifications of negative, trace, and positive. To provide a systematic method of assigning visual readings, the three classifications were defined as follows. Trace was included to allow for uncertainty in visual assessments (a barely visible or questionable test line) as well as to account for disagreement within the schistosomiasis community as to the interpretation of trace results [17]. The test was considered positive if both the test and control lines appeared, with either the test line appearing darker, similar in intensity, or fainter than the control line. And the test result was considered negative when only the control line appeared.

For batches of five tests at the commercially-suggested reading time, POC-CCA readings were entered in a SchistoTrack electronic survey. The survey was programmed in the Open Data Kit (ODK) platform that was accessed by technicians using the ODK Collect application version 2022.4 on Lenovo TB-8505F tablets operated with Android 9 [36]. Importantly, the ODK survey prompted technicians to capture a photo of the POC-CCA cassette. To help control for variation in lighting due to the weather, technicians placed the cassette in the centre of a white plastic lightbox (Linghuang brand, made of polyvinyl chloride) with dimensions 24x23x22cm, lit with an attachable white LED light. Technicians were set up in an area avoiding direct, strong sunlight. A photo of the cassette was taken through the lightbox hole opening (24 cm above the cassette, the height of the lightbox) using the 5MP rear camera of the Android tablet and without zoom. The resulting images were 1920x2560 pixels in dimension with a screen resolution of 72 dots per inch. The ODK survey was configured to automatically save the time at which the photo was taken. Survey entries for each participant, which consisted of the POC-CCA result, a photo of the POC-CCA test, and the photo-taking time, were uploaded daily to a private ODK server at Oxford for later analysis. Any missing results were followed up by the SchistoTrack team. Among the 3224 participants that attended the 2023 SchistoTrack timepoint, a total of 3188 participants provided a valid urine sample and did not have missing information in the ODK survey for urine testing (e.g. due to missing this clinical station within SchistoTrack) and therefore were the focus of this analysis. For participants without missing test results, there were no invalid tests or invalid/removed pictures.

### Senior technician rereading of field images

Following fieldwork, in March 2023 at the Uganda Ministry of Health lab in Kampala, one senior technician with 10 years of experience with POC-CCA tests reread a random sample of 10% of the SchistoTrack field POC-CCA images. The Oxford study team selected, with uniform random probability sampling, 318 participants (with one image each; approx. 10%) and provided a PDF of these images labelled with image IDs. The senior technician was given an Excel sheet (Microsoft Excel, Version 2412, Microsoft corporation) with drop-down lists to record the POC-CCA results for each image ID. The commercially-suggested reading time, which was written by hand on the POC-CCA cassette in the field as stated above, also was recorded by the senior technician. This time entry provided data to compare the commercially-suggested reading time at which the test should have been read with the actual time at which the photo was taken (from the ODK survey). During rereading, the senior technician was blinded to the original readings from the field.

### Reference libraries

We used calibrated *S. mansoni* antigen samples with known concentrations of adult-worm-antigen trichloroacetic acid-soluble fraction AWA-TCA containing CCA, to construct several reference libraries of images and readings of POC-CCA cassettes. Lyophilised samples were produced by Mondial Diagnostics (MondialDx) [37]. AWA-TCA calibrated samples of 11 CCA concentrations ranging from 0.3 to 240 ng/mL (0.3, 1.8, 2.1, 2.4, 3, 4.5, 9, 24, 60, 120, and 240 ng/mL) were used to create reference libraries.

Reference libraries were prepared across three different country laboratories with varying settings and equipment access. All labs used the same POC-CCA batch as was used during the fieldwork. There was an industry lab at MondialDx, an in-country government lab at the Uganda Ministry of Health in Kampala, and a research lab at Vanderbilt University. At each laboratory, the lyophilised samples were rehydrated by adding 500 *µ*L of distilled clean water into each vial, standing the vial for one minute to allow the sample to reconstitute, and mixing gently until all dissolved.

Each laboratory ran every calibrated sample concentration in triplicate. In December 2022, at the MondialDx laboratory, POC-CCA test images were taken at 20 minutes against a plain white background. A ring light (Falcon Eyes Ringlamp FLC-28 + TMB-18Z) behind a camera (Canon Powershot SC620 HS, 20.2MP)was used to take photos at a distance of 24cm directly above the cassette.

In March 2023 in the Uganda Ministry of Health lab, images of each cassette were taken at 20 minutes after preparation. The equipment setup was the exact same as that used for the field images with the exception of being in a building with electricity in Kampala. The results were recorded with the same procedure as the field images in an ODK survey.

In June 2023 at the research lab at Vanderbilt University, a lateral flow reader (LFR) was used to detect the light intensity across the test strip in millivolts (mV) and the test and control line signal peak areas in mm*mV. The lateral flow reader was considered to be the gold-standard measurement of signal intensity for all reference libraries and a benchmark for the linescan pipeline performance. The LFR used was a Qiagen ESEQuant (QIAGEN Lake Constance GmbH, Stokach, Germany). Additionally, time-lapse images were taken of POC-CCA cassettes prepared with AWA-TCA calibrated samples of 2.1, 3, 4.5, and 9 ng/mL CCA concentrations. The series of AWA-TCA calibrated samples were selected to represent a range of concentrations that also includes the assumed commercial detectable limit (2.4 ng/mL antigen concentration) and that might vary with extended light exposure (hence, extremely high concentrations were not included). The cassettes were prepared in triplicate and images were taken once per minute for 60 minutes, starting immediately after preparation, using a camera on a timer at a height of 24cm above the cassettes against a white background. The camera used for time-lapse images was a iPhone 13 Pro, 12MP rear-facing camera and the time-lapse photos were systematically collected at exact intervals of one minute by using the Lens Buddy application.

### Image analysis pipeline

Figure 6 shows a schematic diagram of the automated image analysis method, which extracts the test-to-control ratio from an input image of a POC-CCA test. The method comprises three main stages in the following order (a) a deep learning model for instance segmentation of the buffer well and cassette membrane; (b) reorientation and refinement of the detected membrane; and (c) horizontal line-scanning of pixel intensity and integration of test and control peak areas for finding the test-to-control ratio (Figure 6).

**Figure 6:**
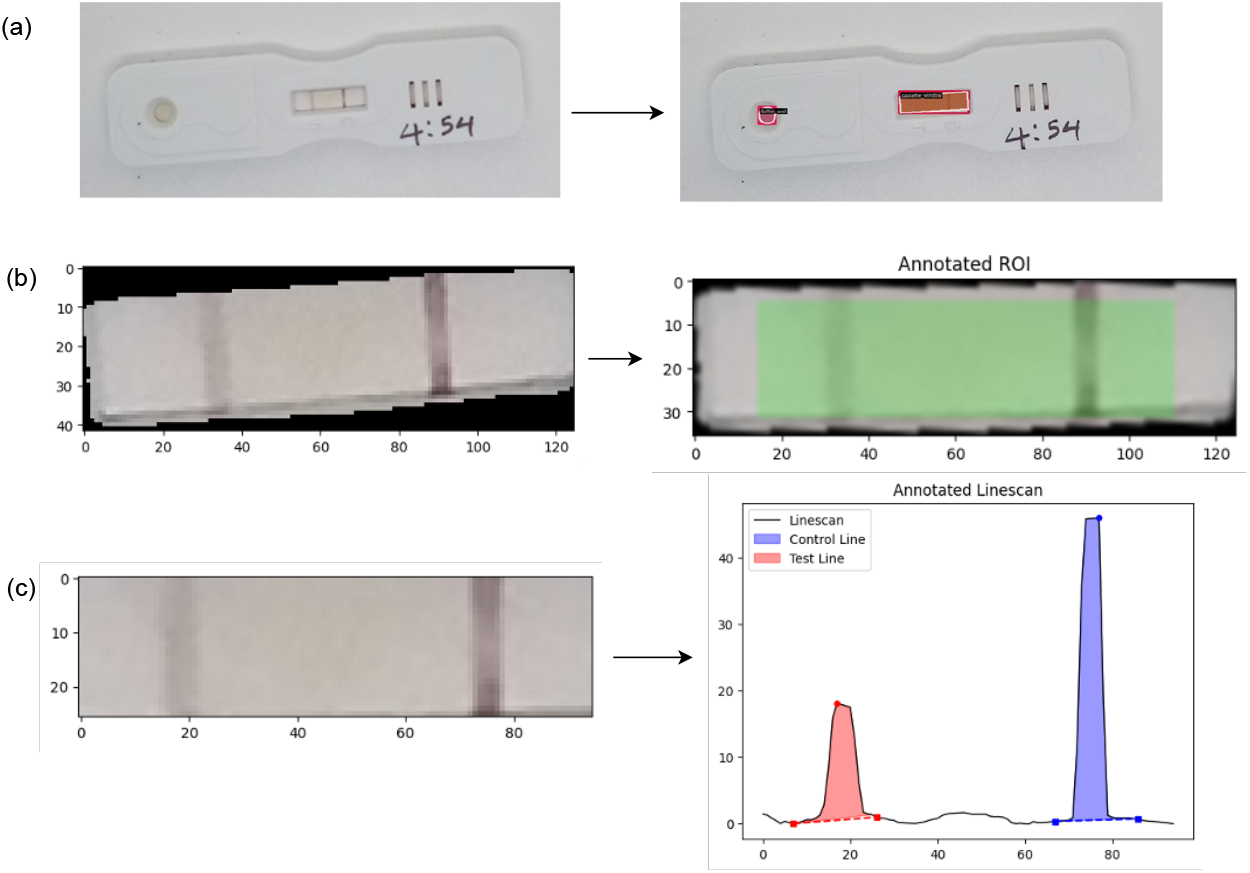
Image analysis pipeline for automated signal processing of lateral flow assays

### Deep learning instance segmentation model

The region-based convolutional neural network model Mask R-CNN was used for instance segmentation of the buffer well and cassette membrane (Figure 6 (a)). Mask R-CNN performs highly on several well-established instance segmentation benchmarks, and has been widely used across several domains including medical imaging, remote sensing, and robotics [38, 39, 40, 41]. We used the implementation from OpenMMLab of Mask R-CNN through their MMDetection framework, version 3.2.0 [42]. The model we used had a ResNet-50-FPN backbone and was pretrained on the Microsoft COCO dataset [43, 44, 45]. We also compared segmentation of the buffer well and cassette membrane to segmentation of the membrane only. We trained two Mask R-CNN models to detect the cassette membrane only, implementing the same ResNet-50-FPN backbone and another with a larger ResNeXt-101-32x8d-FPN backbone [46].

SchistoTrack field POC-CCA images were used for model training and evaluation. We randomly selected 322 field images (independently of images selected for senior technician re-reading) and split into train, validation, and test datasets of sizes 258, 32, and 32, respectively, representing an 80-10-10% split. All 322 images were annotated with the cassette membrane and buffer well using the VIA annotator tool, with polygon and circle annotations used for the membrane and buffer well respectively [47]. Model training on the 258 images was performed for 12 epochs with a batch size of two using stochastic gradient descent for parameter optimisation. The base learning rate (LR) for the optimiser was 0.0025, a momentum factor of 0.9 was used, and the L2 regularisation coefficient was 0.0001. We employed a LR scheduler with a linear warm-up for 500 iterations (approx. four epochs), with the LR linearly increasing from a starting point of 0.001 times the base LR. After linear warm-up, a multi-step LR scheduler was used, where the LR remained constant at the base LR before being decayed stepwise by a factor of 0.1 after 8 and 11 epochs were complete. The validation set of 32 images was used to evaluate the model every three epochs, assessing mean Average Precision (mAP) computed across multiple intersection over union thresholds ranging from 0.50 to 0.95 for the cassette membrane and buffer well classes. The best performing model in terms of mAP was saved as the final model. The final model was then evaluated for COCO mAP on the out-of-sample test set of 32 images. Alongside point estimates of the test set mAP, 95% confidence intervals (CIs) were estimated using bootstrapping with 1000 resamples of the test set. All training and evaluation was performed on a single NVIDIA Tesla V100 GPU with CUDA 11.3.1 installed.

### Reorientation and refinement of detected membrane

Image processing methods were performed using OpenCV-Python v4.7.0 with Python v3.9.6. The segmentation mask of the detected cassette membrane was used to extract the membrane from the image. As the cassettes may not always be photographed in the same position, reorientation of the membrane was performed by first calculating the angle between the line connecting the centres of the detected buffer well and membrane and the horizontal axis, and then using that angle to rotate the membrane such that it was aligned at 0^°^, with the test line on the left and the control line on the right. The reoriented membrane was resized to a standard dimension of 35x135 pixels. The linescan region of interest (ROI) was defined by cropping out a border of five pixels in from each edge as shown by the highlighted area (Figure 6 (b)). The resulting membrane for horizontal line-scanning is shown in Figure 6 (c).

### Line-scans of pixel intensity and peak area integration for test-to-control ratios

The main outcome to be produced from image analysis of the lateral flow assay was the test-to-control ratio, which was extracted from all field images of POC-CCA. It was calculated as the ratio of the test peak area to the control peak area. The POC-CCA test result was deemed positive from the image analysis method if the test-to-control ratio was greater than or equal to 0.15, otherwise the test was considered negative as previously described and validated in Scherr et al [48]. No classifications were defined apriori to what might be considered a trace in the line scan intensities given that trace is only a visual classification. No threshold calibration was done to avoid data-mining a threshold that fits the visual readings. However, we conducted a number of checks for systematic biases when using a test-to-control ratio. We examined Spearman correlations between the test-to-control ratio and the individual test and control signal intensities, and used a Wilcoxon rank-sum test to compare control signal intensities between tests classified as positive and negative by the linescan threshold. The image analysis method entailed linescans of the pixel intensity in the ROI, identification of peaks of the pixel distributions for the test and control lines, and peak area integration to measure the signal intensity.

Linescans, peak identification, and peak area integration were modified following the pipeline described in Scherr et al. for malaria and adapted recently for schistosomiasis [48] [49]. Horizontal linescans of raw pixel intensity were performed across the scanning ROI at every pixel height in the red, green, and blue channels (R, G, B). For each channel, the set of horizontal scans were averaged vertically over all pixel heights to produce a mean pixel intensity scan across the ROI. A grayscale intensity signal was calculated by averaging over the R, G, B channels and then inverting by subtracting the signal from 255 (such that the troughs became peaks). Signal processing methods were applied to refine the grayscale signal. A median filter with a kernel width of seven pixels was used for smoothing. Any linear trend present in the signal was removed by subtracting the result of a linear least-squares fit. Baseline subtraction to remove noise was performed by estimating the signal baseline using a 3^rd^ order polynomial fit, which was then subtracted from the signal. Python v3.9.6 with SciPy v1.11.3 was used for median filtering and detrending, and PeakUtils v1.3.4 for baseline subtraction.

Peaks present in the signal were detected by computing the numeric first-order derivative, using the PeakUtils package. The control line peak was identified as the peak detected after the midpoint, since the membrane was oriented with the control line on the right and test line on the left. Test peak detection was done by setting an estimated distance of 60 pixels from the control peak and an estimated radius of 20 pixels either side of this location. The test peak was identified as the highest point within this estimated radius. The reason for finding the test peak in this manner instead of using the first-order derivative was that in a truly negative POC-CCA test as well as with any lateral flow assay, there will be no peak in intensity at the test line location. Detection of the control line was used not only to calculate a test-to-control ratio, but also for internal image standardisation and as a further check of quality control for the POC-CCA test. Control lines are not calibrated on the POC-CCA test unlike the test line, and conventionally are used only for testing passage of all reagents through the membrane. Accordingly, we conducted sensitivity analyses to assess any potential result re-classification focusing on the test signal alone. Numerical integration via the trapezium rule was used to find the area under the test and control line peaks, with the resulting signal values being unitless. In a few cases, where small dips in pixel intensity around the test peak resulted in a slightly negative test line area, the overall test-to-control ratio was coded as zero. The width of the definite integrals was set as 10 pixels either side of each peak.

### Evaluation of the image analysis pipeline

To assess the accuracy of the image analysis outcomes and potential to produce quantitative outputs, the image analysis pipeline was applied to the images taken from the reference libraries from the industry lab at MondialDx and government lab at the Uganda Ministry of Health. The test-to-control ratios were compared to intensity readings from the LFR from the research lab at Vanderbilt University.

To assess the agreement of our automated pipeline classifications with conventional visual readings, sensitivity and specificity were shown through confusion matrices. Two versions of the visual result were constructed where trace was considered negative and another version where trace was considered positive. Comparisons were done with the automated pipeline results both with the field readings and the senior technician re-readings to account for differences in expertise for visual assessment [5]. The distributions of signal intensities for test lines, control lines, and test-to-control ratios across negative, trace, and positive visual readings also were investigated for both the field technicians and the senior technician.

### Assessment of signal intensities over time

We assessed whether deviations from the commercially-suggested reading time affected the signal intensities extracted from the image analysis pipeline. Field conditions could affect the pixel intensity of the lateral flow assay if there was a delay in the time the image was taken resulting in extended analyte exposure to air and light. We evaluated whether the amount of time in minutes past the optimal (as commercially suggested) reading time of 20 minutes affected the signal intensities extracted from the image analysis pipeline. The intensities of the test line, control line, and test-to-control ratio were correlated against the difference in minutes between the commercially-suggested reading time and image timestamp from ODK. These associations were further examined by subgroups of the visual reading assignment by field technicians of negative, trace, and positive. The field technician assignments were studied in case there was an association as this could have affected the signal present in the image that was reread by the senior technician. The infection burden of the individual might affect the analyte concentration and therefore, the test-to-control ratios or relation of signal intensities over time. Eggs per gram (EPG) of stool were measured through Kato-Katz (KK) microscopy as described elsewhere [33]. Test line, control line, and test-to-control ratio intensities against the commercially-suggested reading time also were re-examined by sub-groups of WHO categories of *S. mansoni* infection burden [12] of none, light (1-99 EPG), moderate (100-399 EPG), and heavy (400 EPG). To understand if disagreement between the image analysis outcomes and the visual readings was due to deviations from the commercially-suggested reading time, the median number of minutes for images in agreement was compared against images with disagreement. The distribution of signal intensities was plotted for all images in disagreement to assess whether there was variability in the image analysis outcome or the visual reading assignment. Agreement also was reassessed by removing any images with a time difference *>* 20 minutes or incorrectly recorded time difference of *<*-20.

To ascertain the time-to-positivity, including before the commercially suggested reading time, as well as whether there could be any increase or decay in signal intensities over extended periods of time under controlled conditions, the time lapse reference library from the research lab at Vanderbilt University was analysed with the image analysis pipeline in June 2023. Signal intensities were plotted at one-minute intervals from sample preparation up to 60 minutes, i.e. up to 40 minutes after the commercially-suggested reading time, for each replicate.

### Comparison to deep learning classification

We compared the performance of our signal processing pipeline with commonly used deep learning models, including ResNet-50 [50], MobileNetV2 [51], and a vision transformer (ViT) [52] for classifying POC-CCA test results [4]. These classifiers were trained, validated, and tested on labeled ROIs which were extracted and processed from 3188 field images following steps (a) and (b) of our image analysis pipeline (Figure 6). To compare models trained with the two different visual assignments, two separate classification experiments were conducted: one experiment with visual labels assigned by field technicians and another experiment with labels from the re-reading of 318 field images by a senior technician. For both cases, the visual labels were binarised in two ways, with trace treated as positive in one and negative in the other. For all experiments, the labeled datasets were split into training, validation, and test sets using a 70-15-15% stratified split, resulting in dataset sizes of 2230, 479, and 479 for the field-based labels, and 222, 48, and 48 for the senior technician labels. Larger validation and test splits (15 as opposed to 10) than those used for segmentation was implemented given the fewer labels from the senior technicians. The performance of each classifier was evaluated using sensitivity, specificity, and accuracy metrics on the test set associated with each experiment. In our study, we considered the visual assignments of 318 field images by the senior technicians to be the closest approximation to the ground truth for deep learning classification, as the reader had more experience than field technicians, and these labels are based on images rather than direct field observations of the cassettes or other patient characteristics. Thus, in a final experiment, the classifiers trained on the pre-segmented images labeled by the senior technician were used to classify the field-labeled ROIs, excluding the ROIs of the 318 re-read images. For comparison to the performance of linescans, sensitivity, specificity, and accuracy also were calculated for the linescan-based classification results on the corresponding test sets of each experiment.

All three deep learning classifiers were implemented in PyTorch and initialised with weights pretrained on the ImageNet [53] dataset. Training was performed using a batch size of eight and the SGD optimiser with a momentum of 0.9 and L2 weight decay of 0.0001. The LR was set to a maximum value of 0.0001 and adjusted dynamically using a OneCycle LR scheduler, which gradually increased the LR from an initial value set to 1/25 of the maximum LR during a warm-up phase lasting approximately two epochs, followed by cycling and decay of the LR over the remainder of the training with a minimum possible value of 1/10^4^ of the maximum LR. Each model was trained on an NVIDIA A100 GPU and evaluated on the respective validation set at every epoch. To prevent overfitting, early stopping was employed based on the average validation accuracy over a rolling 10-epoch window. If no improvement in the average validation accuracy by a minimum value of 0.0001 was observed within a period of 10 epochs, training was halted and the model with the highest rolling average was saved. The total training times were recorded for each experiment. All images were resized to 384x384 pixels before being input to the models. Data augmentation, including random brightness and contrast jittering, small affine translations, and Gaussian blurring, was applied during training. These augmentations were implemented using the torchvision.transforms library module in Python, with parameters set to brightness=0.5 and contrast=0.5 for color jittering, translate=(0.01, 0.01) for random affine transformations, and kernel size=3 with sigma=(0.1, 1.0) for Gaussian blurring. Final evaluation was conducted on the test datasets using sensitivity, specificity, and accuracy metrics. Alongside point estimates of these metrics, 95% CIs were estimated using bootstrapping with 1000 resamples of the test set.

### Quantitative classifications from test-to-control ratios

We explored the potential for quantitative diagnoses from our automated pipeline. In addition to comparing the linescans to the known concentrations in reference samples, which investigates the ability to produce a sensible quantitative output, we also examined the association of the test-to-control ratio, and the signal intensities of the test and control lines, against the natural log of EPG plus one (from KK microscopy), and against two previously used semi-quantitative visual rankings using 1) artifically printed POC-CCA tests (also called G-scores [54]) that provide a means of standardising visual assessments of line intensities and 2) comparisons of test-to-control lines to create positivity rankings of pos1 to pos3. Detailed descriptions are provided in the Supplementary Methods.

## Data Availability

Image data are not available due to restrictions in ethics approvals and agreements with participants as well as to the ongoing nature of the SchistoTrack Cohort. Model code has been provided as supplementary material. Code specific to the line scanning only is available upon reasonable request to TFS.

## Declarations

### Author contributions

Conceptualisation: GFC. Data curation: CH, CP, BN, CPM, RA, RB, KB, JO, RK, TS, and GFC. Formal analysis: CH, CP. Funding acquisition: G.F.C. Investigation: CH, CP, and GFC. Methodology: CH, CP, TS, and GFC. Project administration: BN, NBK, and GFC. Resources: RP, NBK, GVD, GFC. Software: GFC. Supervision: TS and GFC. Validation: TA, BN, CH, CP, CPM, TS, and GFC. Visualisation: CH and CP. Writing - original draft: CH, CP, and GFC. Writing - review & editing: CH, CP, BN, PN, CPM, TA, RP, PTH, NBK, RB, KB, JO, RK, GVD, TS, GFC.

### Funding

Grants from the Wellcome Trust Institutional Strategic Support Fund (204826/Z/16/Z), NDPH Pump Priming Fund, and Robertson Foundation Fellowship, and UKRI EPSRC (EP/X021793/1) were funded to GFC. This research was funded in whole, or in part, by the UKRI (EP/X021793/1). For the purpose of Open Access, the author has applied a CC-BY public copyright licence to any Author Accepted Manuscript version arising from this submission. TPS and CPM were supported by the United States National Institutes of Health (R01AI163472).

### Declaration of interests

TA and RP were employed by MondialDx. GVD occasionally acts as a consultant to MondialDx. All other authors declare no competing interests.

## Acknowledgments

We are thankful to the SchistoTrack teams in both Oxford and Uganda for their feedback, survey help, data collection, and constant support throughout this project. Dr. Lauren Wilburn provided useful feedback in investigating quality control of future POC-CCA batches. Special thanks go to the district teams in Uganda and local political leadership as well as the community medicine distributors. We are grateful for the constant involvement through the study timepoints and community engagement meetings of our study participants.

## Ethics approvals

Data collection and use were reviewed and approved by Oxford Tropical Research Ethics Committee (OxTREC 509-21), Vector Control Division Research Ethics Committee of the Uganda Ministry of Health (VCDREC146), and Uganda National Council of Science and Technology (UNCST HS 1664ES). Written informed consent was obtained from adult participants aged 18 years and older, who also provided written consent on behalf of verbally assented children with fingerprint assent. Older children also provided written consent in addition to the adult consent on their behalf.

