## Supplementary material for "Interpretable machine learning and signal processing for automated reading and quality control of lateral flow tests for schistosomiasis"

### Supplementary information

#### Supplementary methods

##### Semi-quantitative visual assessments

For direct visual assessment used to produce a semi-quantitative reading, subjective comparative intensities of control and treatment lines were used. The visual classification was as follows: negative if only the control line appeared; trace if the test line was barely visible; positive if both the test and control lines appeared.

The mock set of cassettes, henceforth called G-scores [1] was produced and donated by Leiden University Medical Centre (Batch 202212). G-scores have been proposed as a method to standardise visual assessments among technicians. There was a set of 10 G-score cassettes that were printed with a range of linearly increasing pixel intensities for printed treatment lines (G1-G10) and one set pixel intensity for the control line. Technicians lined up the ten printed G-scores cassettes in order from G1-G10, and matched the participant POC-CCA test and control lines to the most visually similar G-score test line, and recorded the reading. Though, the original printing of the G-scores was calibrated to represent various intensities of test lines, not considering the exact variation anticipated in control lines.

For both sets of visual readings, the assigned G-score of the test line was divided by the visually assigned G-score of the control line to construct G-score ratios for an additional comparison to the extracted field test-to-control ratios. As a robustness check, since printed G-score intensities were intended to match test lines, we also compared G-scores of the test line with extracted test signals.

#### Supplementary tables

Table S1: Summary of signal intensity values. Median test and control line signal intensities overall and by each visual category assigned by technicians in the field, with sample size $n$, difference in medians, and Wilcoxon signed-rank test statistics.

| **Visual result** | **Test signal** | **Control signal** | $n$ | **Difference** | $W$ | $p$**-value** |
|---|---|---|---|---|---|---|
| All | 24.9 | 200.4 | 3188 | 175.5 | 833 | $< 0.05$ |
| Negative | 6 | 171.4 | 992 | 165.4 | 8 | $< 0.05$ |
| Trace | 15.2 | 196.5 | 529 | 181.3 | 0 | $< 0.05$ |
| Positive | 79 | 220.2 | 1667 | 141.2 | 693 | $< 0.05$ |

Table S2: Comparison of distribution fits for test-to-control ratios. BIC values for lognormal, gamma, and exponential distribution fits to the test-to-control ratio overall and for different categories of visual assignment.

| Category | Lognormal | Gamma | Exponential |
|---|---|---|---|
| Overall | -56.15 | 222.51 | 7805.94 |
| Negative | -1914.72 | -1337.36 | 2085.50 |
| Positive | -53.50 | -29.04 | 4519.13 |
| Negative + trace | -3301.89 | -2535.50 | 3224.54 |
| Positive + trace | 84.69 | 152.20 | 5687.13 |

Table S3: Classifier performance comparisons.

| Classifier | Trace treatment | Trained on senior re-readings and evaluated on field readings | | |
|---|---|---|---|---|
| | | Sensitivity | Specificity | Accuracy |
| ResNet50 | Positive | 0.550 [0.527, 0.573] | 0.964 [0.951, 0.976] | 0.681 [0.663, 0.699] |
| MobileNetV2 | Positive | 0.535 [0.513, 0.55] | 0.967 [0.955, 0.978] | 0.672 [0.655, 0.689] |
| ViT | Positive | 0.727 [0.707, 0.746] | 0.991 [0.984, 0.997] | 0.811 [0.797, 0.825] |
| Linescan | Positive | 0.676 [0.657, 0.696] | 0.985 [0.976, 0.992] | 0.774 [0.759, 0.790] |
| ResNet50 | Negative | 0.660 [0.636, 0.682] | 0.996 [0.992, 0.999] | 0.821 [0.807, 0.834] |
| MobileNetV2 | Negative | 0.658 [0.634, 0.681] | 0.992 [0.987, 0.997] | 0.818 [0.804, 0.832] |
| ViT | Negative | 0.763 [0.741, 0.785] | 0.984 [0.977, 0.990] | 0.869 [0.857, 0.882] |
| Linescan | Negative | 0.864 [0.847, 0.882] | 0.964 [0.955, 0.974] | 0.912 [0.901, 0.923] |

### Supplementary figures

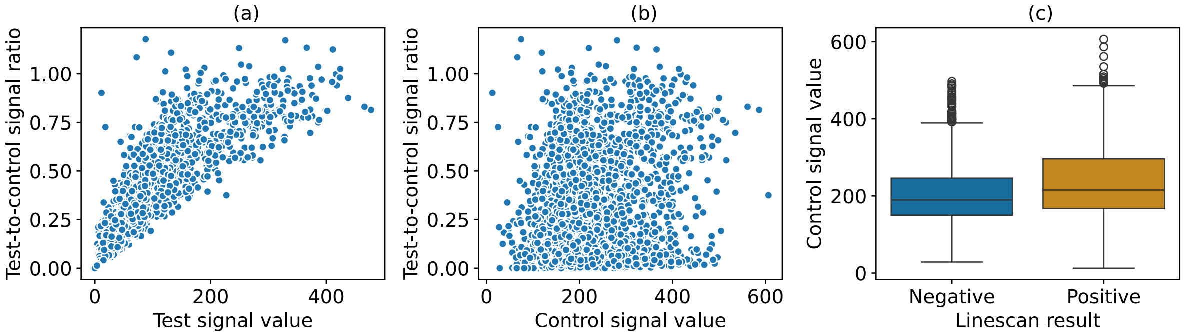


Figure S1: Checks for systematic bias. The figure shows scatter plots of the test-to-control ratio against the test signal intensity (a), and the control signal intensity (b), and a box plot of the control signal values classified by linescan results (c).

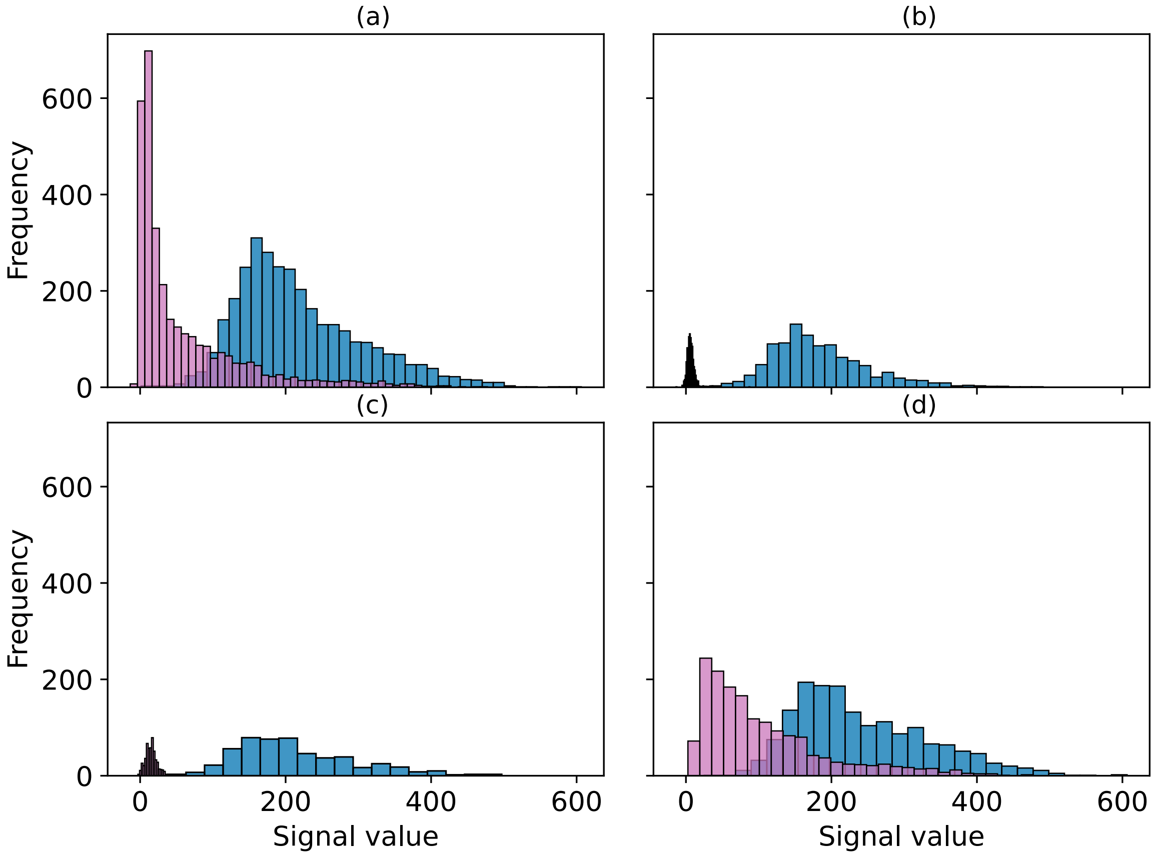


Figure S2: Distribution of test (pink) and control (blue) line signals overall (a), and categorised by technician-assigned test result (negative (b), trace (c), positive (d)).

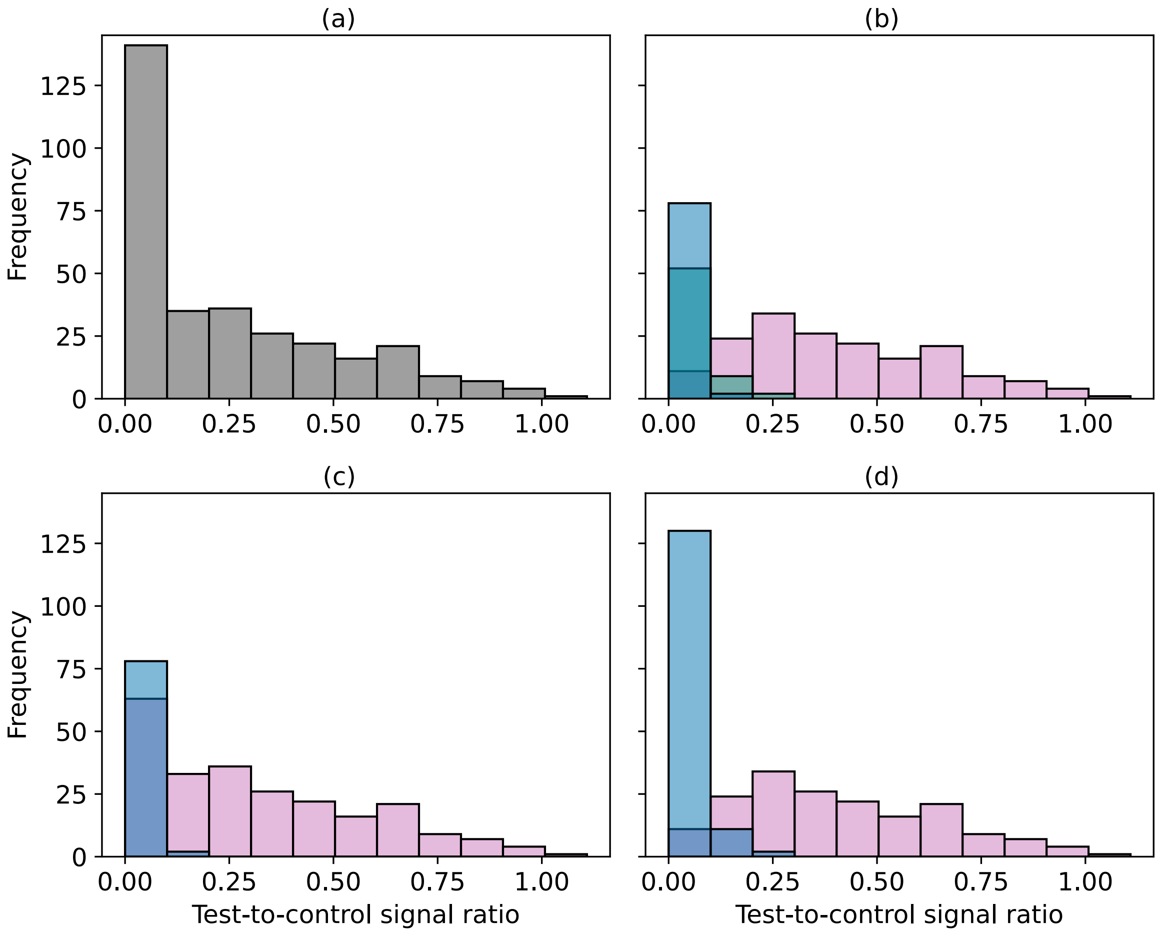


Figure S3: Distribution of test-to-control ratios for 318 images reread by senior technician based on visual assignments by field technicians. (a) Overall distribution, (b) Distribution by positive (pink), trace (green), and negative (blue), (c) Distribution by positive (pink) and negative (blue) where trace is considered positive, and (d) Distribution when trace is considered negative.

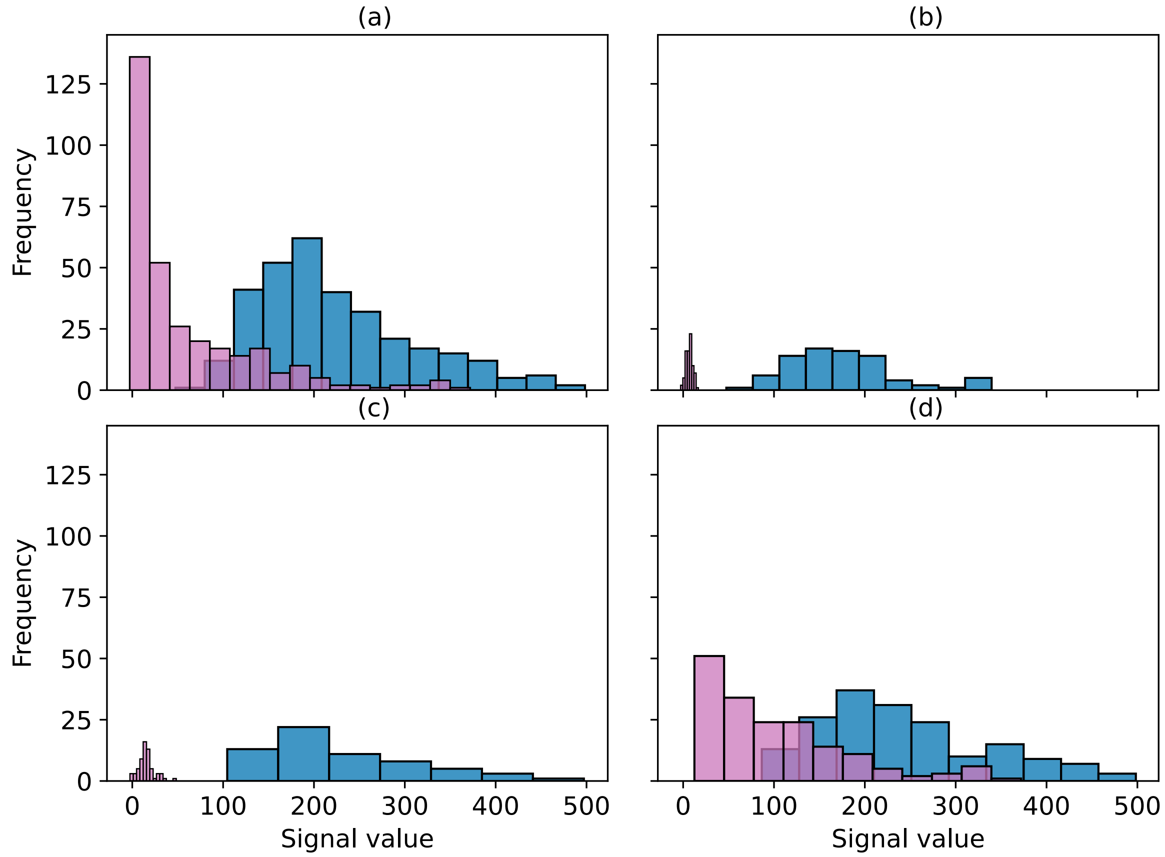


Figure S4: Distribution of test (pink) and control (blue) line signals for images reread by senior technician, overall (a), and categorised by field technician-assigned test result (negative (b), trace (c), positive (d)).

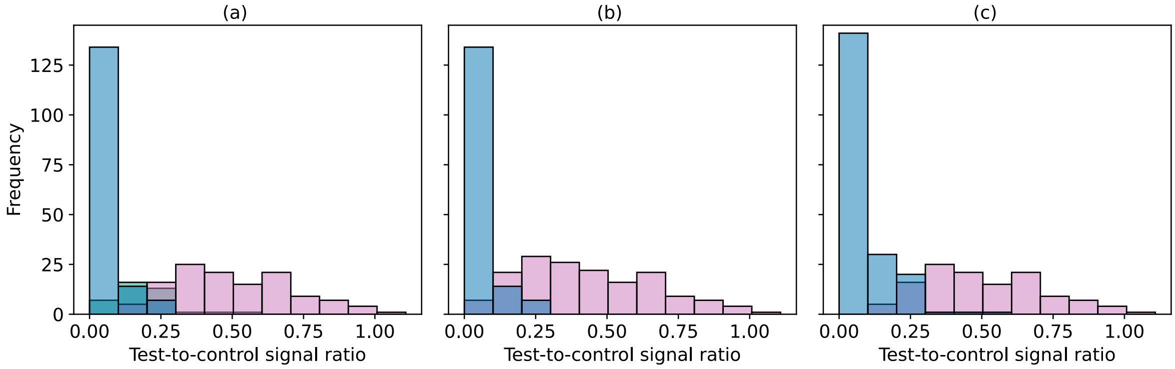


Figure S5: Distribution of test-to-control ratios for 318 images reread by senior technician based on their visual assignments. (a) Distribution by positive (pink), trace (green), and negative (blue), (b) Distribution by positive (pink) and negative (blue) where trace is considered positive, and (c) Distribution when trace is considered negative.

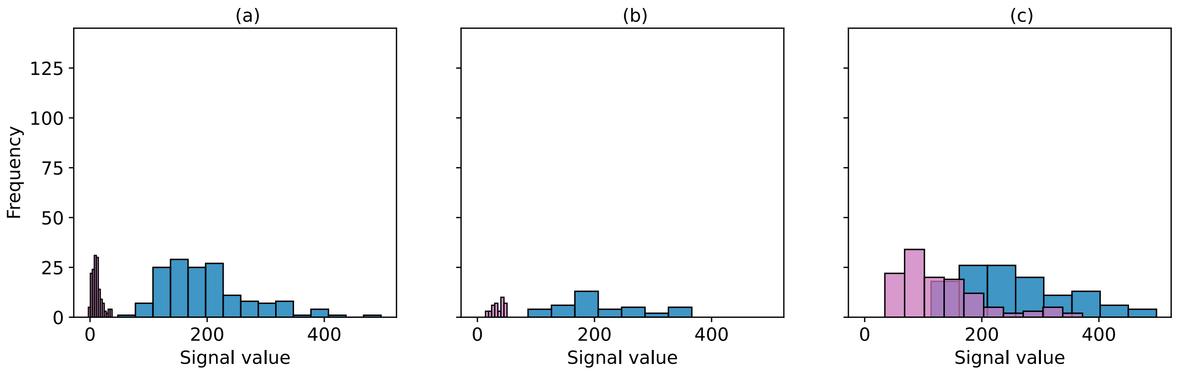


Figure S6: Distribution of test (pink) and control (blue) line signals for images reread by senior technician, categorised by test result assigned by them (negative (a), trace (b), positive (c)).

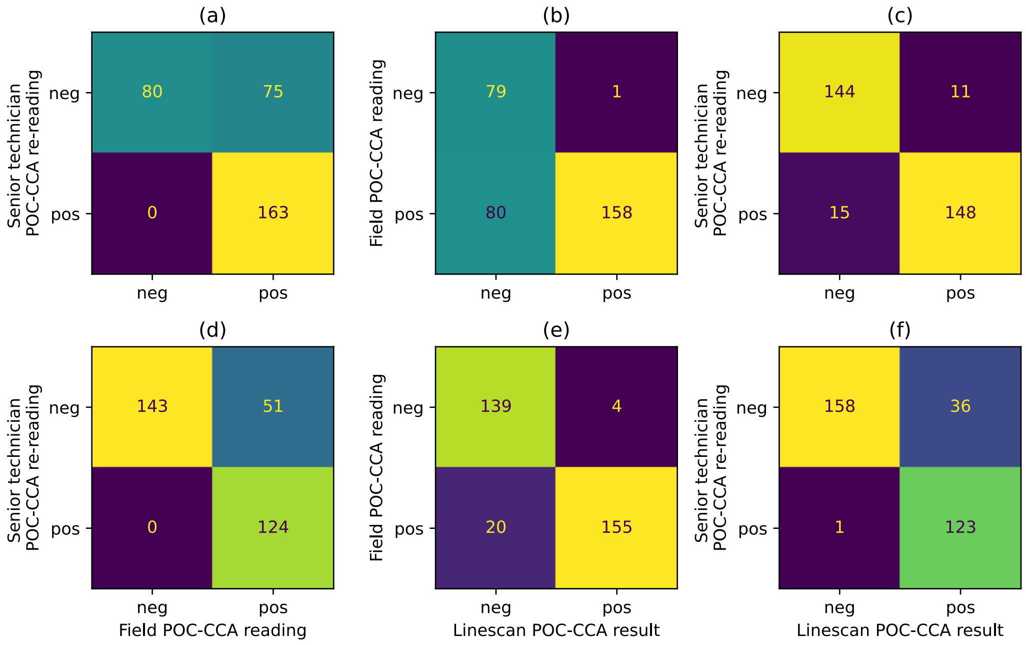


Figure S7: Confusion matrices of agreement among visual readings and image analysis outcomes. (a) shows the agreement between field technician and senior technician (sensitivity=100% (163/163), specificity=51.6% (80/155)), (b) the agreement between field technician and image analysis outcome (sensitivity=66.4% (158/238), specificity=98.7% (79/80)), and (c) the agreement between senior technician and image analysis outcome (sensitivity=90.8% (148/163), specificity=92.9% (144/155)). (d)-(f) are repeated in the same order as (a)-(c) but with trace recoded from positive to negative for visual readings. The sensitivity and specificity values are 100% (124/124) and 73.7% (143/194) for the confusion matrix in (d), 88.6% (155/175) and 97.2% (139/143) for the one in (e), and 99.2% (123/124) and 81.4% (158/194) for the one in (f).

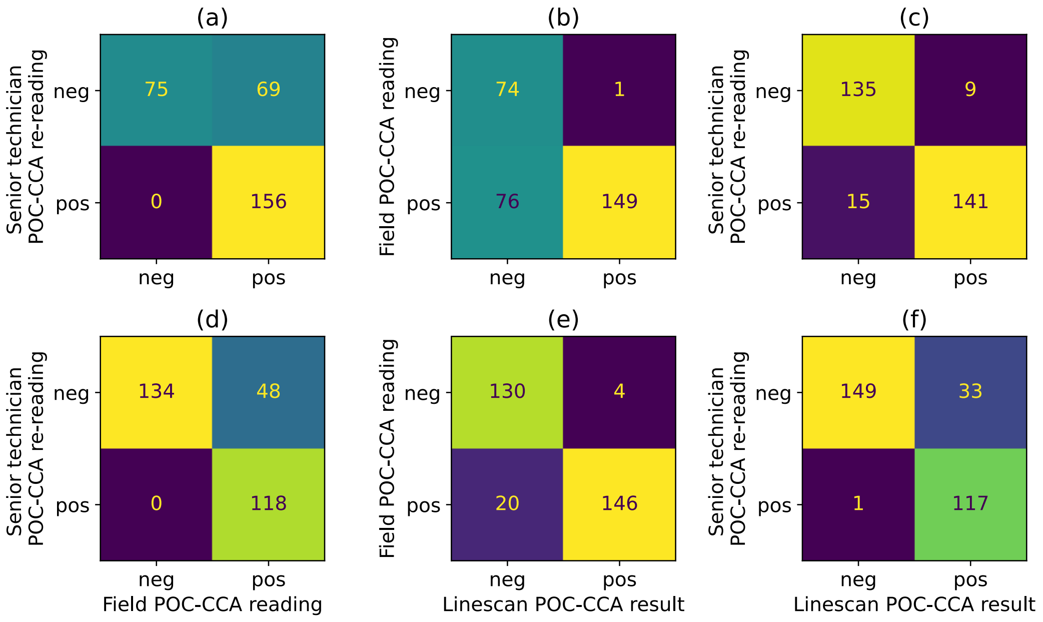


Figure S8: Confusion matrices of visual reading and linescan agreement after excluding invalid images based on reading time. The total number of images considered here was 300, after excluding 18 images, of which one image had an incorrectly recorded time difference of $<$-20 minutes, and 17 images had time differences $>$ 20 minutes. (a) shows the agreement between field technician and senior technician (sensitivity=100% (156/156), specificity=52.1% (75/144)), (b) the agreement between field technician and image analysis outcome (sensitivity=66.2% (149/225), specificity=98.7% (74/75)), and (c) the agreement between senior technician and image analysis outcome (sensitivity=90.4% (141/156), specificity=93.8% (135/144)). (d)-(f) are repeated in the same order as (a)-(c) but with trace recoded from positive to negative for visual readings. The sensitivity and specificity values are 100% (118/118) and 73.6% (134/182) for the confusion matrix in (d), 88% (146/166) and 97% (130/134) for the one in (e), and 99.2% (117/118) and 81.9% (149/182) for the one in (f).

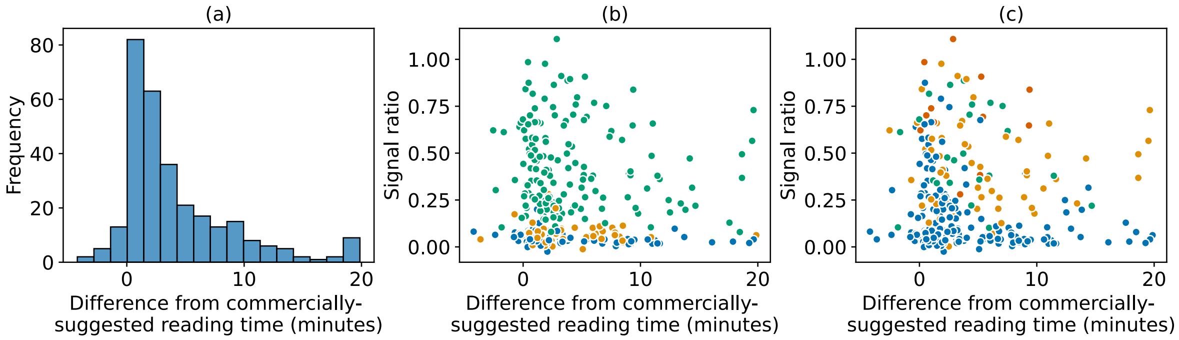


Figure S9: Time differences from the commercially-suggested reading time. A subset of 300/318 field images re-read by a senior technician is shown. There were 18 images excluded from the plots due to outlier values for the time difference. One image had an incorrectly recorded time difference of <-20 minutes, and 17 images had time differences > 20 minutes. (a) Distribution of time differences for images re-read by the senior technician. (b) Scatter of test-to-control ratio against time difference. No association was observed overall (Spearman's $\rho$ = -0.05, p = 0.39). The colors represent the visual reading categories from the field technician. Negative (blue), trace (orange), or positive (green). Spearman correlation between test-to-control ratio and time difference was statistically significant only for visually negative results (Spearman's $\rho$ = -0.35, $p < 0.05$). No significant correlation was observed for trace ($\rho$ = -0.01, p = 0.91), or positive ($\rho$ = -0.04, p = 0.57) classifications. (c) Scatter of test-to-control ratio against time difference, with colors representing the WHO infection intensity categories from KK microscopy. None (blue), light (orange), moderate (green), or heavy (red). Spearman correlation between test-to-control ratio and time difference was statistically significant only for the no infection category (Spearman's $\rho$ = -0.23, $p < 0.05$). No significant correlation was observed for light ($\rho$ = -0.03, p = 0.83), moderate ($\rho$ = -0.02, p = 0.94), or heavy ($\rho$ = -0.19, p = 0.57) intensity categories.

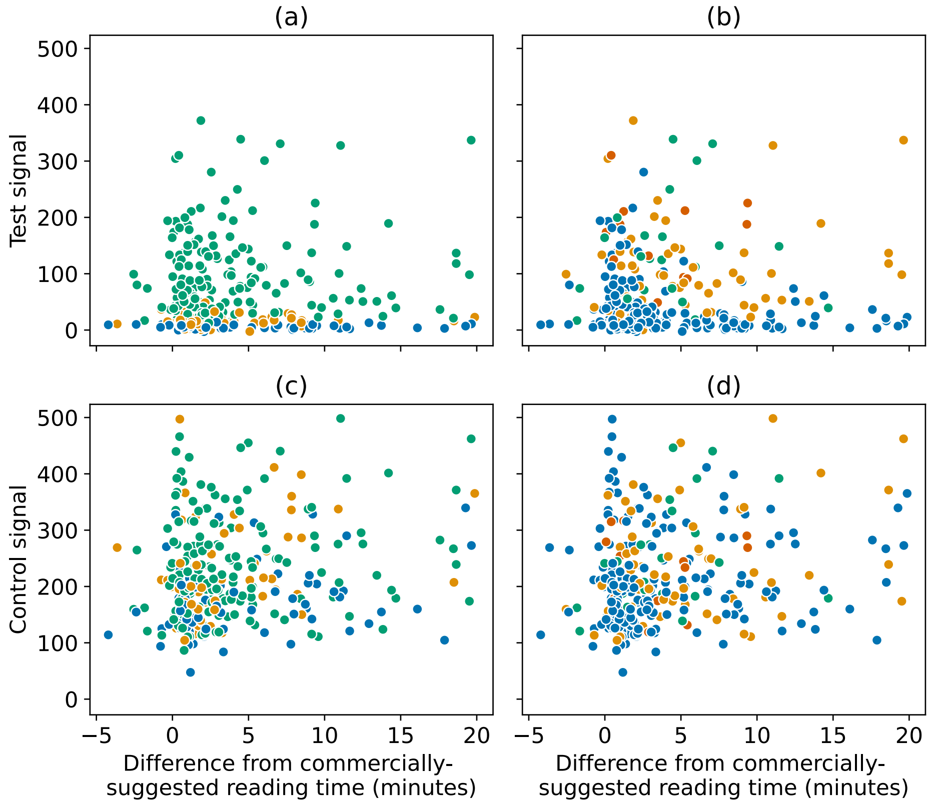


Figure S10: Test and control signal values against time difference. A subset of 300/318 field images re-read by a senior technician are shown. There were 18 images of which one image had an incorrectly recorded time difference of <-20 minutes, and 17 images had time differences > 20 minutes, excluded from the plots. (a) Scatter of test signal values against time difference, with colors representing the visual reading categories from the field technician. No association was observed overall (Spearman's $\rho = 0$, p = 0.99). Negative (blue), trace (orange), or positive (green). Spearman correlation between test-to-control ratio and time difference was statistically significant only for visually negative results (Spearman's $\rho$ = -0.26, $p < 0.05$). No significant correlation was observed for trace ($\rho = 0.18$, p = 0.16), or positive ($\rho = 0.07$, p = 0.35) classifications. (b) Scatter of test signal values against time difference, with colors representing the WHO infection intensity categories from KK microscopy. None (blue), light (orange), moderate (green), or heavy (red). Spearman correlation between test-to-control ratio and time difference was statistically significant only for the no infection category (Spearman's $\rho$ = -0.17, $p < 0.05$). No significant correlation was observed for light ($\rho = 0.14$, p = 0.28), moderate ($\rho = 0.17$, p = 0.44), or heavy ($\rho = 0.15$, p = 0.63) intensity categories. (c) Scatter of control signal values against time difference, with colors representing the visual reading categories from the field technician. Negative (blue), trace (orange), or positive (green). Spearman correlation between test-to-control ratio and time difference was statistically significant overall (Spearman's $\rho = 0.15$, $p < 0.05$) and for visually trace results (Spearman's $\rho = 0.34$, $p < 0.05$), but insignificant for negative ($\rho = 0.13$, p = 0.23), or positive ($\rho = 0.13$, p = 0.09) classifications. (d) Scatter of control signal values against time difference, with colors representing the WHO infection intensity categories from KK microscopy. None (blue), light (orange), moderate (green), or heavy (red). Spearman correlation between test-to-control ratio and time difference was statistically significant only for the moderate infection category (Spearman's $\rho = 0.43$, $p < 0.05$). No significant correlation was observed for no infection ($\rho = 0.08$, p = 0.25), light ($\rho = 0.2$, p = 0.12), or heavy ($\rho = 0.05$, p = 0.87) intensity categories.

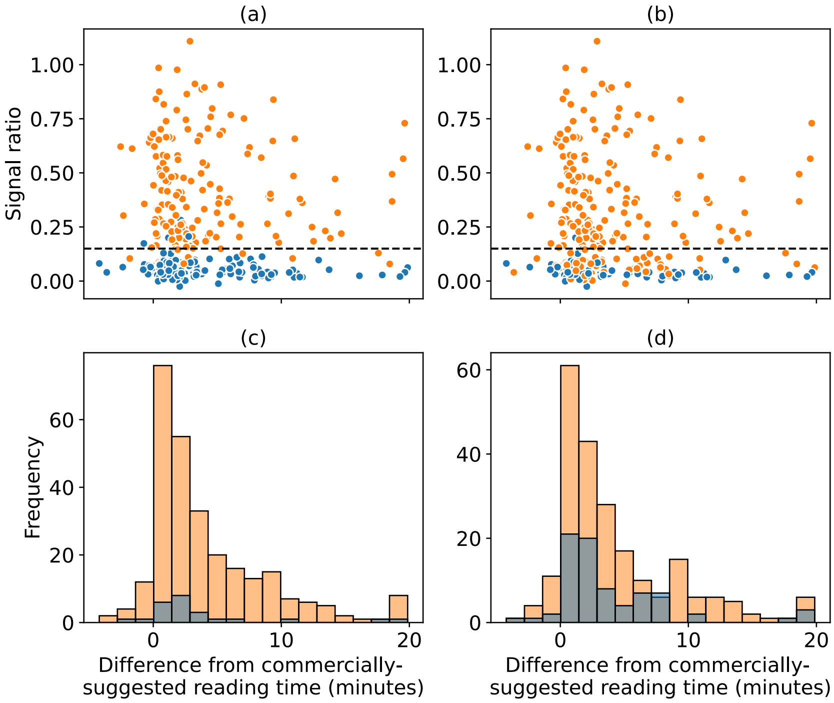


Figure S11: Reading time difference based on agreement between linescan and field results. The test-to-control ratios with respect to difference from the commercially-suggested reading time and the distribution of the time difference for a subset of 300/318 field images re-read by a senior technician are shown. There were 18 images of which one image had an incorrectly recorded time difference of <-20 minutes, and 17 images had time differences > 20 minutes, excluded from the plots. (a) Scatter of test-to-control ratio against time difference, with colors representing the visual reading categories from the field technician when the trace is considered negative. Negative (blue), or positive (orange). (b) Same as (a), but with trace considered positive. (c) Distribution of time difference for images in agreement (orange), and disagreement (blue), when trace is considered negative. (d) Distribution of time difference for images in agreement (orange), and disagreement (blue), when trace is considered positive.

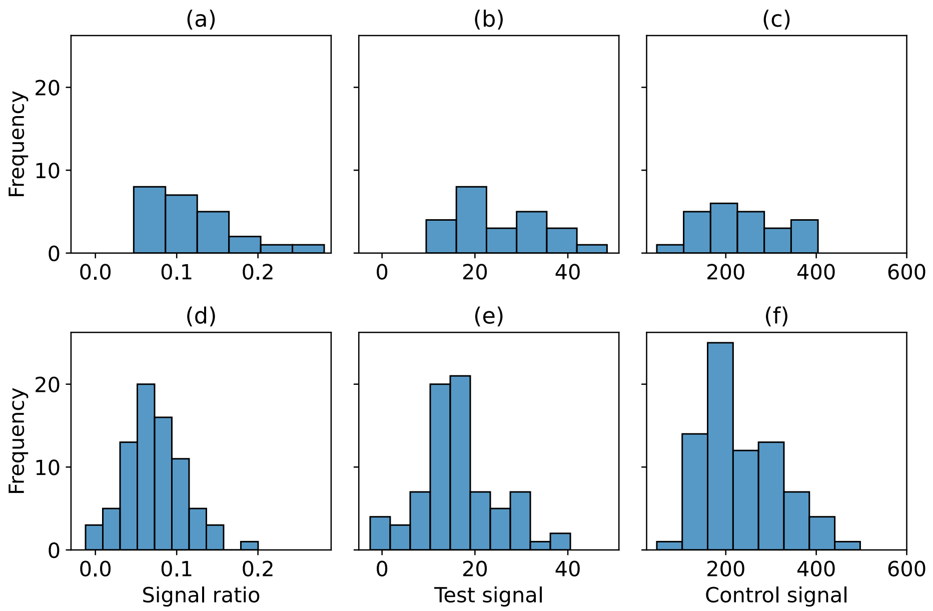


Figure S12: Distribution of signals for disagreed results. The figure shows the distribution of the test-to-control ratio, test signal and control signal for images with disagreement between the field and the linescan result. (a-c) show the distributions for 24 field images in disagreement with linescan results when visual trace reading is considered negative. (d-f) show distributions of 77 field images in disagreement with linescan results when visual trace results are considered positive.

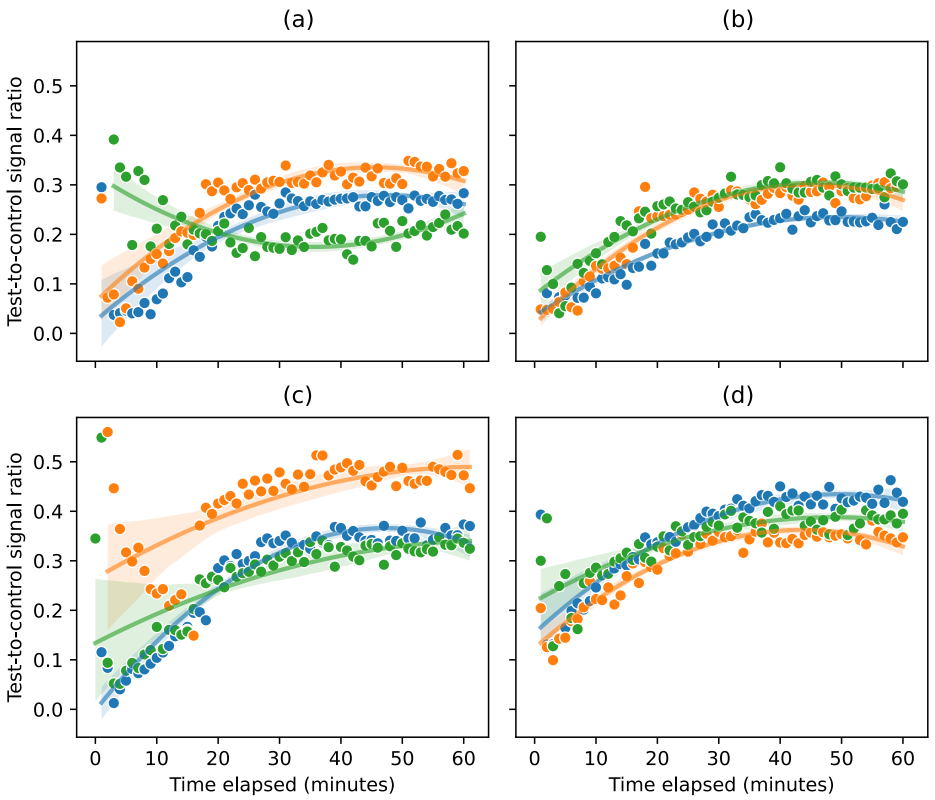


Figure S13: Time-to-positivity. Polynomial fits are shown for each replicate (orange, blue, green each representing a different replicate). (a) 2.1 ng/mL CCA calibrated sample. The unusual trend in one replicate (green) in the figure was driven by observing no change in the test line over time (see Supplementary Figure S14(e)). (b) 3 ng/mL CCA calibrated sample. (c) 4.5 ng/mL calibrated sample, (d) 9 ng/mL calibrated sample.

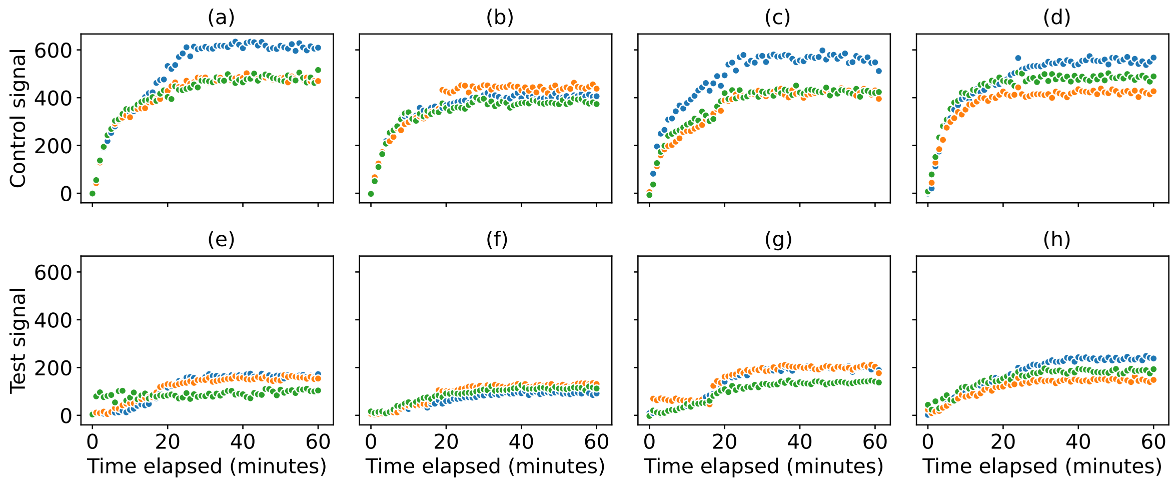


Figure S14: Time-to-positivity for test lines and appearance of control lines. Images of the cassettes were taken once per minute immediately after POC-CCA test preparation for a total of 60 minutes. (a)-(d) Control line signal peak area over time for calibrated samples of CCA concentrations 2.1 ng/mL, 3 ng/mL, 4.5 ng/mL, 9 ng/mL. (e)-(h) Test line signal peak area over time, for calibrated samples with 2.1 ng/mL CCA, 3 ng/mL CCA, 4.5 ng/mL CCA, 9 ng/mL CCA.

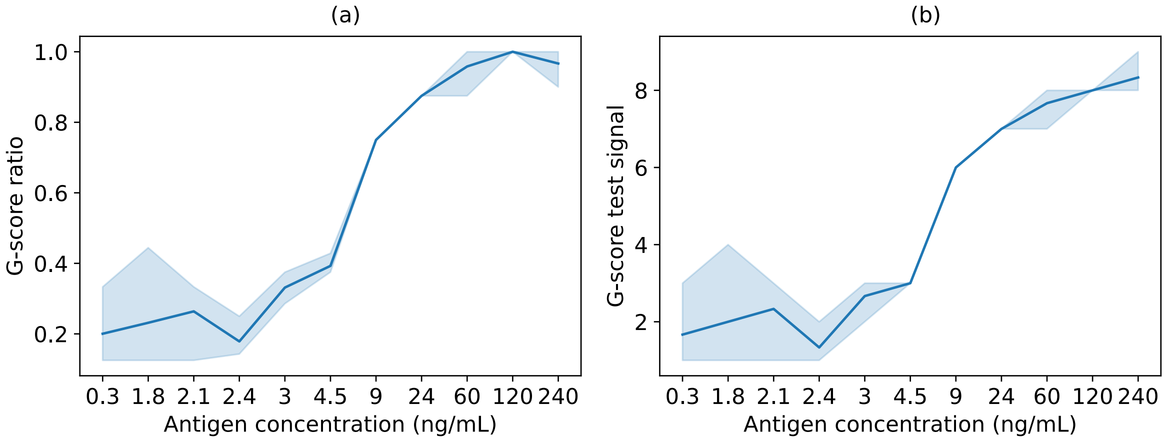


Figure S15: G-score ratios and test line G-scores for the reference library. G-score ratios and test line G-scores for the AWA-TCA calibrated sample series reference library prepared at the government lab at the Uganda Ministry of Health are shown. The graph plots the G-score ratios (a) and the test line G-scores (b) for the 11 calibrated samples examined. The shading shows a 95% CI derived from a bootstrap distribution of the three replicates, resampled 1000 times. The solid lines indicates the mean of the three replicates.
